# Identification of distinct and shared biomarker panels in different manifestations of cerebral small vessel disease through proteomic profiling

**DOI:** 10.1101/2024.06.10.24308599

**Authors:** Ines Hristovska, Alexa Pichet Binette, Atul Kumar, Malin Wennström, Chris Gaiteri, Linda Karlsson, Olof Strandberg, Shorena Janelidze, Danielle van Westen, Erik Stomrud, Sebastian Palmqvist, Rik Ossenkoppele, Niklas Mattsson-Carlgren, Jacob W Vogel, Oskar Hansson

## Abstract

The pathophysiology underlying various manifestations of cerebral small vessel disease (cSVD) remains obscure. Using high-throughput proteomics, we identified common and distinct proteomic signatures of white matter lesions (WML), microbleeds, infarcts and their subtypes, measured in 1,670 living patients. Across all cSVD manifestations, proteins indicative of extracellular matrix dysregulation and vascular remodeling were elevated, including ELN, POSTN, CCN2 and especially MMP12, implicating endothelial and smooth muscle cells of the brain. These proteins were validated in CSF from two additional datasets, and a subset detected in plasma predicted future cerebrovascular events in the UK Biobank better than risk scores currently used in clinical practice. Analysis focusing on WMLs found microglial-associated proteins to be associated with faster WML progression, whereas specific neuron-derived proteins mediated the link between WMLs and longitudinal cognitive decline. These data provide a comprehensive atlas of cSVD biomarkers, and our findings provide a promising roadmap for future diagnostics and therapeutics.

## INTRODUCTION

Manifestations of cerebral small vessel disease (cSVD) have emerged as the leading vascular contributors to cognitive decline and subsequent dementia^1^, and as major risk factors for stroke^2^. cSVD can act as both a primary etiology and as a secondary pathology in numerous neurodegenerative conditions. These manifestations stem from a variety of pathological processes that affect the brain’s small arteries, arterioles, venules, and capillaries, leading to structural and functional abnormalities within the brain’s microvascular network^3^. Although lifestyle changes and controlling vascular risk factors can reduce the risk for cSVD^4^, the development of more targeted preventative and therapeutic strategies is impeded due to the relatively limited understanding of the molecular pathophysiology of cSVD.

At present, clinical detection of cSVD relies strongly on magnetic resonance imaging (MRI), which identifies markers such as white matter lesions (WML), subcortical infarcts and microbleeds^5^. Neuropathological studies and model systems have helped to characterize isolated pathways contributing to the etiology of cSVD^6,7,8^, but the larger landscape and timeline of altered pathways in living humans has been elusive. While existing literature offers insights into pathophysiological mechanisms for cSVD, including endothelial dysfunction, BBB breakdown and inflammation, these findings have not been consistently replicated, requiring further validation across larger cohorts with a broader spectrum of markers^9,10^. Associative evidence exists linking certain lesions together and with other diseases, but ultimately, we have little insight into their underlying etiology^11^. In this context, it is also important to understand how these cSVD-specific changes compare to broader cerebrovascular disease (CVD), such as those associated with large vessel disease^12^. Additionally, MRI lesions related to cSVD pathology are likely indicative of later stages in the pathological processes, underscoring the necessity of investigating the early changes that may precede these visible abnormalities. Such gaps in our knowledge emphasize the need for investigation using richer biomarker panels and larger cohorts to provide deeper insight into the pathophysiological processes involved in cSVD. Discovering robust markers and defining molecules related to earliest stages of disease will be crucial to the development and monitoring of therapeutic interventions.

Recent progress in highly sensitive and high throughput protein measurement techniques now allow for a comprehensive large-scale proteomic profiling in cerebrospinal fluid (CSF) and blood^13–15^. These advances enable identification of novel biomarkers and delineate molecular mechanisms associated with disease pathogenesis *in vivo*^16,17^. In our study, we utilized the proximity extension assay (Olink) proteomics technology to extensively assess pathophysiological pathways concurrent with cSVD manifestations in CSF of several large population and patient cohorts. Our principal aims were i) to identify both common and unique CSF proteome signatures associated with WML, microbleeds and infarcts, and compare profiles according to etiology; ii) to provide in-depth characterization of the biological underpinnings of those signatures; iii) to identify proteins implicated in WML progression and mediating WML-associated cognitive decline, iv) to validate novel biomarkers in CSF and plasma cohorts, and v) to assess their utility in plasma in predicting future cerebrovascular events in a large independent population-based study.

## RESULTS

We analyzed cerebrospinal fluid (CSF) protein levels for 2,943 proteins in 1,670 participants from the Swedish BioFINDER-2 cohort, ranging from cognitively unimpaired individuals to patients with mild cognitive impairment and dementia (Extended Data Fig. 1). Participants were stratified into groups reflecting the presence or absence of CVD, namely WML, microbleeds or infarcts. We distinguished 1,113 participants without WML pathology and 557 with WML pathology (Supplementary Fig. 1). Similarly, individuals were classified based on the presence of microbleeds (1350 without and 269 with ≥ 1 microbleed(s)) and infarcts (1445 without and 225 with ≥1 infarct(s)). Extended Data Table 1 indicates associations between the presence of each CVD pathology and various demographic and clinical measures. As expected, participants with CVD manifestations tended to be older, more likely male, more likely demented and were more likely diagnosed with vascular risk factors compared to subjects without CVD.

### Existence of shared and distinct proteomic signatures in different CVD manifestations

We first identified differentially abundant proteins (DAP) related to broader cerebrovascular disease in individuals exhibiting WML pathology (n=707 proteins), microbleeds (n=84), or infarcts (n=228, pFDR<0.05, Fig. 1a). Almost all associations remained significant after adjusting for co-morbid Alzheimer’s or Parkinson’s disease (Extended Data Fig. 2). Since we aimed to identify and study proteins specifically associated with each of the three CVD manifestations, proteins were retained for further analysis only if their association with WML pathology, microbleeds, or infarcts, respectively, persisted when adjusting for the other two manifestations of CVD (e.g., proteins associated with WML when adjusting for presence of microbleeds and infarcts). Many proteins remained significant after adjusting for other CVDs (Fig. 1b). Relative to microbleeds and infarcts, a large part of the measured proteome exhibited alterations in the presence of WML. Summary statistics from the differential expression analyses can be found in Supplementary Table 1.

**Figure 1.**
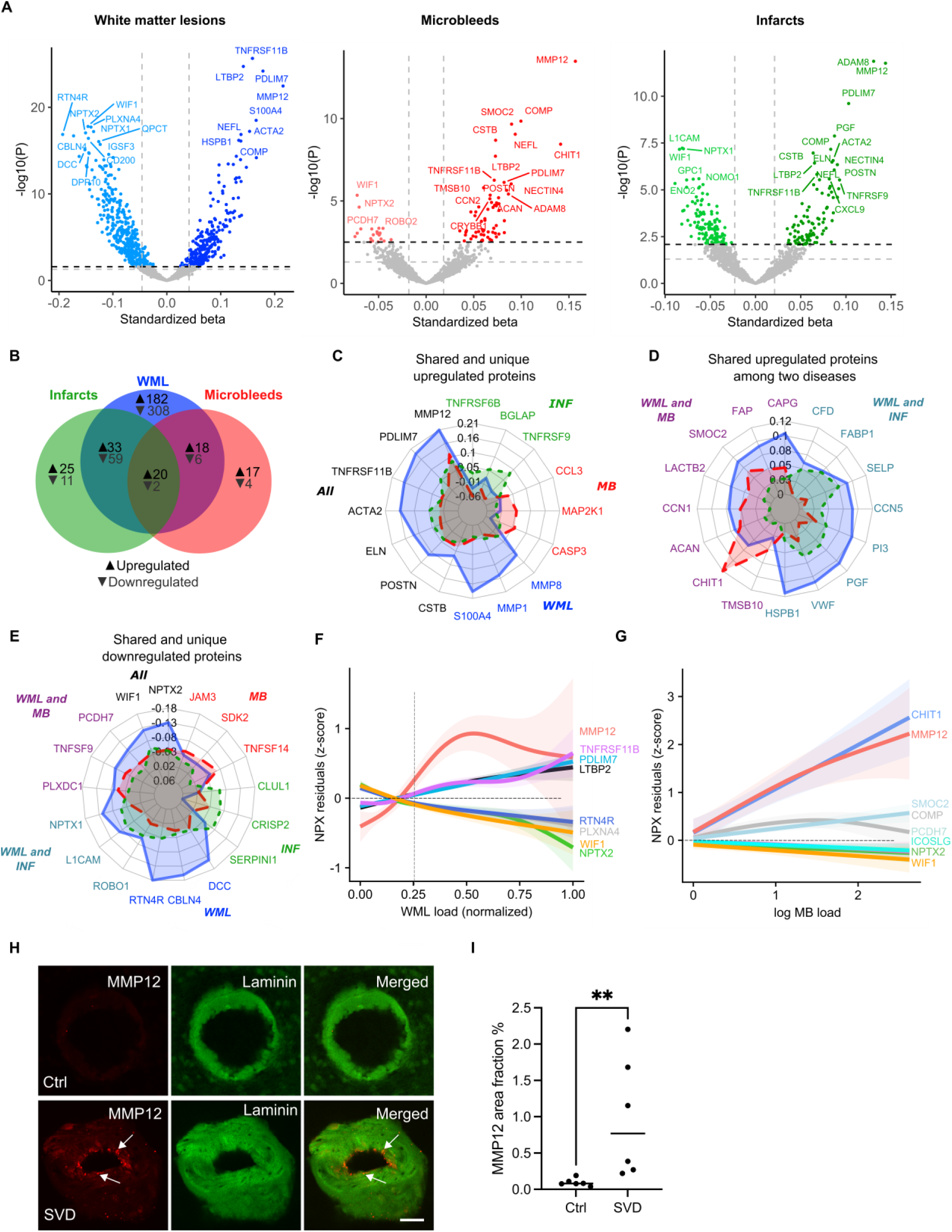
Differential protein expression in subjects with WML, microbleeds, and infarcts. **(a)** Volcano plots showing differentially abundant proteins (DAP) when comparing individuals with and without specific cSVD pathology: WML, microbleeds, and infarcts. The models are adjusted for age, sex, and average protein level. The dashed lines represent significance threshold at ɑ=0.05 before (gray) and after (black) FDR correction. Proteins below the p[FDR]<0.05 threshold were considered significant. For clarity, only the top 20 proteins are labeled. **(b)** Venn diagram illustrating the shared and exclusive proteins associated with WML, microbleeds, and infarcts, with proteins of increased abundance indicated in black and those with decreased abundance in gray. **(c-e)** Radar charts depicting standardized beta coefficients for top proteins, in relation to their association to disease: **(c)** shared and disease-specific upregulated proteins; **(d)** proteins commonly upregulated between two conditions; and **(e)** shared and unique downregulated proteins. Each axis represents a different protein, with axis length indicating the standardized beta coefficients magnitude for each condition. (**f-g)** Proteins of interest from panel **(a)**, selected based on disease relevance or expression pattern, plotted against normalized WML load **(f)** and microbleed burden **(g)**. Shaded areas correspond to the 95% confidence interval. The vertical line indicates the cutoff for WML positivity. **(h)** Representative images showing MMP12 (red, indicated by arrows) within laminin-positive (green) small arteries in control and SVD cases in the frontal lobe white matter tissue. **(i)** Area fraction of MMP12 in arteries of six controls and six SVD patients. Each point represents the mean MMP12 area fraction within laminin-stained regions from five arteries per case. Data was analyzed using the Mann-Whitney U test; **p<0.01. Scale bars = 50 µm.

Several proteins, including MMP12, POSTN and ELN, showed a significant *increase* in abundance across all three manifestations examined of CVD independently (Fig. 1c). Interestingly, these proteins form a tightly connected cluster predominantly associated with extracellular matrix organization (Supplementary Fig. 2). However, the expression of many proteins was found to be specific to a certain cSVD manifestation, most notably in WML. For instance, matrix metalloproteinases such as MMP1 and MMP8, were uniquely elevated in WML. CCL3, involved in immune response, and CASP3, a key player in apoptosis, were specifically increased in microbleeds. On the other hand, several tumor necrosis factor receptor superfamily members, including TNFRSF9 and TNFRSF6B, were upregulated only in infarcts. This differential protein expression pattern with little shared upregulation between microbleeds and infarcts is in agreement with the hypothesis that these manifestations may have distinct underlying etiologies. However, some proteins were found to be increased in WML as well as in either microbleeds or infarcts, as illustrated in Fig. 1d. A similar pattern was observed for proteins that were *downregulated*, with WIF1, involved in Wnt signaling and NPTX2, involved in synaptic function, consistently decreased across all forms of CVD. Compared to microbleeds and infarcts, many proteins were downregulated in the context of WML, with CBLN4 and RTN4R showing substantial negative associations (Fig. 1e).

We next sought to investigate how key proteins fluctuated in response to continuous (as opposed to binary) measures of WML (Fig. 1f) and microbleed (Fig. 1g) loads, respectively. Among the proteins that showed a gradual decrease, NPTX2 exhibited the most notable reduction as the severity of WML increased. MMP12 levels rose sharply in both conditions as pathology became more pronounced, with CHIT1 being more specific with increasing load of microbleeds.

Given the prominent role of MMP12 in cSVD, with strong associations observed across WML, infarcts and microbleeds, we further explored its expression using postmortem data (Fig. 1h). Our analysis revealed elevated MMP12 expression within laminin-rich areas of small arteries in cSVD patients compared to controls, strengthening the evidence of its involvement in the vascular pathology of cSVD (Fig. 1i).

### Stratification of cerebrovascular disease based on topography reveals distinct proteomic profiles

Building on our initial findings, we next stratified the data to investigate whether distinct proteomic signatures correspond to different topographies. Specifically, given that the topography of microbleeds is considered to reflect their underlying etiology, with deep microbleeds more associated with cSVD and lobar microbleeds with amyloid-β/CAA, we explored whether variations in their locations were mirrored by specific protein expression profiles (Fig. 2a-2b). Our analysis identified distinct proteomic profiles associated with either deep or lobar microbleeds (Fig. 2a, Extended Data Fig. 3a). While most proteins were specific to one type, a few, such as MMP12, CSTB, and COMP, were common to both deep and lobar microbleeds (Fig. 2b). However, these shared proteins were also significant in WML and infarcts, indicating they are not specific to microbleeds alone. Notably, CCL3, ITGB2 and GLOD4 were unique to lobar microbleeds, whereas THBS4, RNASE3 and BGLAP were exclusive to deep microbleeds. Interestingly, most DAPs related to deep microbleeds overlapped with those associated with WML, and to a lesser extent, infarcts – a pattern not common for lobar microbleeds. Building on the increase in MMP12 and CHIT1 levels with escalating microbleed pathology (Fig. 1g), we investigated whether this pattern differed between lobar and deep microbleeds. Our analysis revealed that MMP12 levels rose in both cases, whereas CHIT1 levels showed an increase comparable to MMP12 specifically with worsening lobar microbleed pathology (Extended Data Fig. 3b).

**Figure 2.**
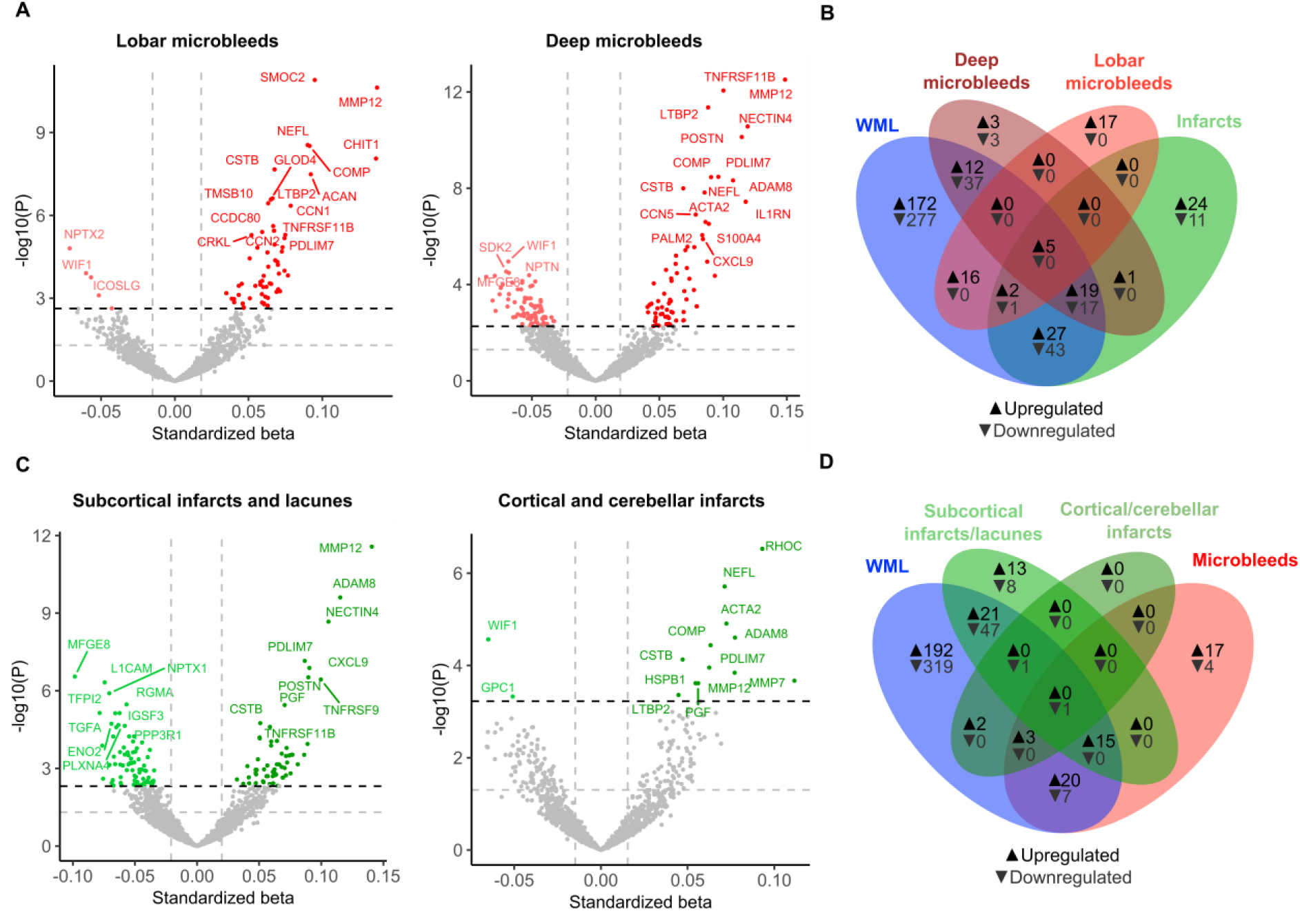
Differential protein expression stratified by cerebrovascular disease topography. **(a)** Volcano plots showing DAP when comparing individuals with and without lobar and deep microbleeds. The models are adjusted for age, sex, and average protein level. The dashed lines represent significance threshold at ɑ=0.05 before (gray) and after (black) FDR correction. Proteins below the p[FDR]<0.05 threshold were considered significant. For clarity, only the top 20 proteins are labeled. **(b)** Venn diagram illustrating the shared and exclusive proteins associated with WML, deep microbleeds, lobar microbleeds and infarcts, with proteins of increased abundance indicated in black and those with decreased abundance in gray. **(c)** Volcano plots showing DAP when comparing individuals with and without subcortical infarcts/lacunes and cortical/cerebellar infarcts. The models are adjusted for age, sex, and average protein level. **(d)** Venn diagram illustrating the shared and exclusive proteins associated with WML, subcortical infarcts/lacunes, cortical/cerebellar infarcts and microbleeds, with proteins of increased abundance indicated in black and those with decreased abundance in gray.

Additionally, we separated infarcts into cortical/cerebellar infarcts, typically linked to large vessel disease, and subcortical infarcts and lacunes, more associated with cSVD (Fig. 2c-2d, Extended Data Fig. 3c). We identified a few significant proteins associated with cortical and cerebellar infarcts, with minimal overlap with subcortical infarcts and lacunes. Notably, RHOC, a key regulator of vascular homeostasis, emerged as a top hit linked to cortical and cerebellar infarcts. In contrast, subcortical infarcts and lacunes exhibited a proteomic profile more closely aligned with WML and, to a lesser extent, deep microbleeds, indicating a stronger association with cSVD. Unique proteins associated with subcortical infarcts and lacunes included those involved in immune regulation and metabolism, such as members of the tumor necrosis factor superfamily, as well as C3, CA1, and CA3. This overlap highlights the closer relationship between deep microbleeds, subcortical infarcts/lacunes, and WML, in contrast to the distinct profile observed in cortical/cerebellar infarcts and lobar microbleeds. Additionally, very few proteins were downregulated in cortical/cerebellar infarcts and lobar microbleeds, further distinguishing them from other SVD-associated manifestations.

### cSVD is associated with greater neurovascular and lesser neuronal cellular responses

To better understand the specific cellular mechanisms underlying the different cSVD manifestations, we sought to uncover the cell-type specificity of DAP and their biological roles. For cell-type specificity, we used Expression Weighted Cell Type Enrichment (EWCE) on single-cell transcriptome data from the SEA-AD atlas and the Human Brain Vascular Atlas, the latter providing greater granularity into vascular cells thought to underpin cSVD.

Proteins *upregulated* across cSVD manifestations, in WML and deep microbleeds, predominantly originated from vascular-associated cells, particularly vascular leptomeningeal cells (VLMC) and smooth muscle cells (SMC) (Fig. 3a). Further dissection of DAP in smooth muscle cell populations showed a significant enrichment specifically in arterial (as opposed to arteriolar) SMC. Interestingly, WML-associated proteins were uniquely enriched in perivascular fibroblasts.

**Figure 3.**
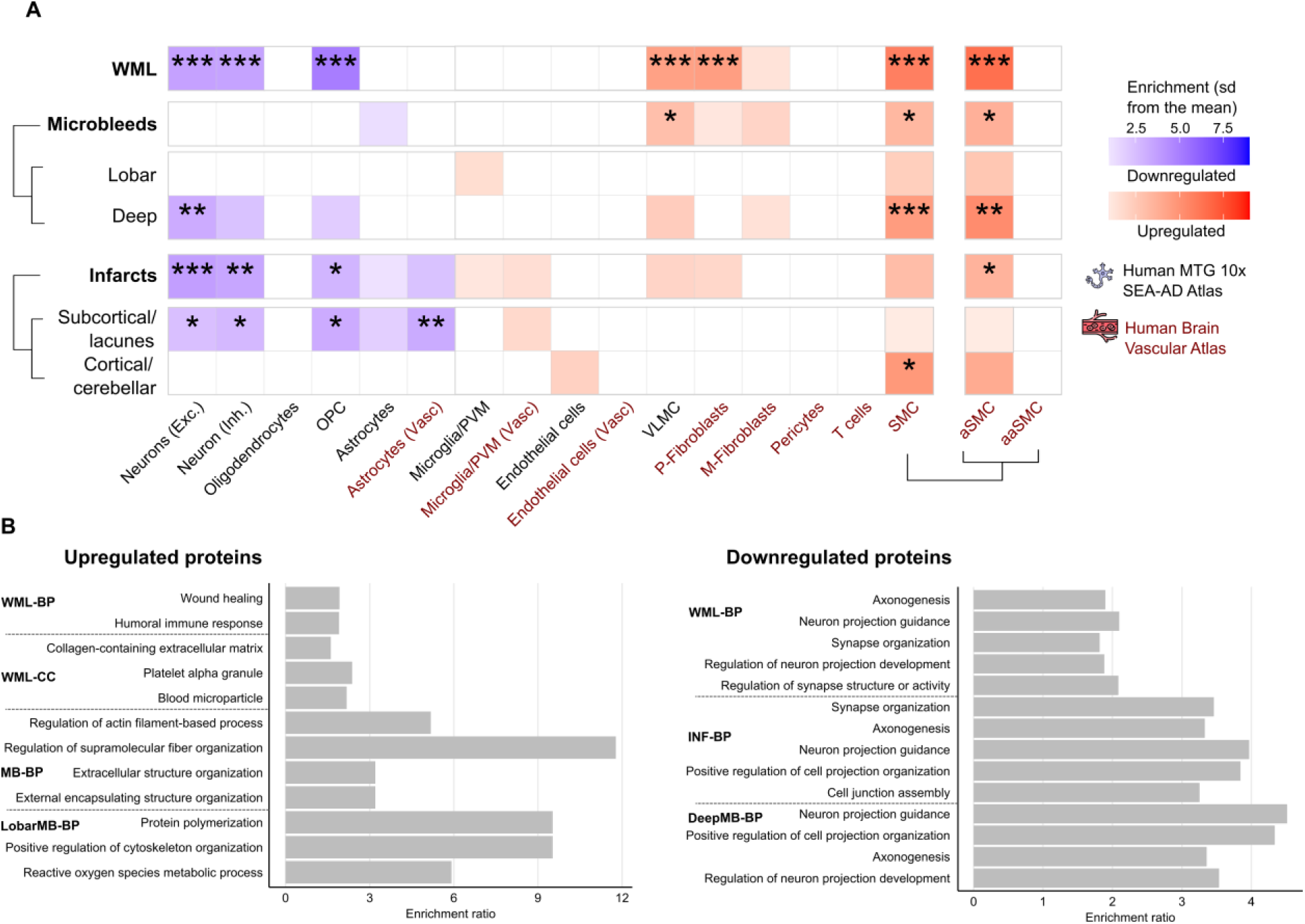
Cell-type and functional enrichment characterization of DAP. **(a)** Cell-type enrichment analyses for upregulated and downregulated proteins based on single-cell transcriptomics data from the Human MTG 10x SEA-AD and the Human Brain Vascular Atlas for different DAP categories using the Expression Weighted Cell Type Enrichment (EWCE) package. The enrichment for upregulated proteins is shown in a palette of red, while the enrichment for downregulated proteins is shown in a palette of blue. **(b)** Top 5 summary terms from functional enrichment analyses using Gene Ontology (GO) databases (Biological Processes and Cellular Components) for different DAP categories. For cell-type and functional enrichment analyses, the 1388 Olink proteins were used as background. *corresponds to p_FDR_<0.05, **corresponds to p_FDR_<0.01, ***corresponds to p_FDR_<0.001. Abbreviations: OPC = oligodendrocyte precursor cells, PVM = perivascular macrophages, VLMC = vascular leptomeningeal cell, P-Fibroblasts=perivascular fibroblasts, M-Fibroblasts = meningeal fibroblasts, SMC = smooth muscle cell, aSMC = arterial smooth muscle cell, aaSMC=arteriolar smooth muscle cell, BP = Biological Processes, CC = Cellular Components, MF = Molecular Functions, IgC = Integral Component, IC = Intrinsic Component, PM = Plasma Membrane

Conversely, *downregulated* proteins were predominantly associated with neuronal cells – both excitatory and inhibitory – as well as oligodendrocyte precursor cells (OPCs) for WML and subcortical infarcts/lacunes, indicating a distinct cellular response within the neuronal and oligodendrocyte populations. A similar trend was observed in deep microbleeds, though with significant enrichment only in excitatory neurons. Notably, only subcortical infarcts/lacunes exhibited significant enrichment of downregulated proteins in astrocytes, highlighting a unique cellular response to this pathology.

Building on these cellular insights, we sought to understand the biological significance of DAP using functional enrichment analysis with the Gene Ontology (GO) database (Fig 3b). All significant GO terms are listed in Supplementary Table 2. Upregulated proteins in WML highlight wound healing, immune responses, and extracellular matrix remodeling, with significant overlap in the latter seen in both WML and microbleeds. Lobar microbleeds were uniquely associated with oxidative stress responses and protein polymerization. Conversely, downregulated proteins across WML, infarcts, and deep microbleeds consistently pointed to disruptions in neuronal processes, such as axonogenesis and synapse organization, suggesting a common underlying neuronal and OPC impairment.

To further explore the molecular mechanisms underlying cSVD, we used the SpeakEasy2^18^ network-based clustering algorithm to identify co-abundance modules in CSF proteomics data, subsequently annotating these modules for cell-type specificity and biological processes (Extended Data Fig. 4a-d). The assignment of proteins to each module is found in Supplementary Table 1. Among the 10 modules analyzed, those with increased protein abundance in SVD were primarily linked to vascular and immune responses (Extended Data Fig. 4e). Notably, Module 5, enriched with proteins involved in metabolism, found in endothelial and arterial smooth muscle cells (e.g. MMP12, PDLIM7, and ACTA2), showed a marked increase with WML and microbleeds even at lower pathology levels, while Module 3, associated with immune functions, steadily rose without plateauing at higher WML and microbleed loads (Extended Data Fig. 4f-g). Modules with decreased protein abundance were primarily associated with neuronal processes, enriched in proteins linked to OPC and inhibitory neurons, and involved in axonogenesis and developmental growth regulation. Additionally, WML was negatively correlated with Module 8, which was enriched in proteins associated with synaptic vesicle organization and transport in both excitatory and inhibitory neurons. Thus, cSVD is associated with alterations in metabolic, vascular, immune and neuronal processes along the cSVD continuum.

### Contribution of immune-related proteins in the progression of WML over time

Our data expectedly showed an overall increase in WML volume over time in subjects with longitudinal follow-up (n=856 subjects, average follow-up time=2.94±1.02, Supplementary Fig. 3a). We proceeded in identifying the proteins at baseline that were associated with the rate of longitudinal increase in WML burden by applying linear mixed-effect models. This analysis revealed that abundance of several proteins at baseline was associated with faster WML progression over time (Fig. 4a, Supplementary Table 4). To gain a deeper insight into the role these proteins play in WML progression, we divided them into two categories: those associated with both baseline and longitudinal WML (shared, in blue), and those associated with longitudinal WML only (longitudinal-only, in magenta). Approximately half of the longitudinal WML-associated proteins were also associated with WML at baseline, indicating sustained upregulation, with NEFL, MMP12 and COMP being prominent among these proteins. In contrast, immune-related proteins, such as ITGB2, and CHI3L1 showed strong associations only with the rate of longitudinal WML progression, but not WML at baseline. In line with this observation, cell-type enrichment analysis indicated that shared proteins were significantly enriched in VLMC (per the SEA-AD Atlas), as well as perivascular fibroblasts and smooth muscle cells (per the Human Vascular Atlas) (Fig. 4b). Conversely, longitudinal-only proteins were significantly enriched in microglial and macrophage populations, suggesting important contribution of immune-related proteins in the progression of WMLs.

**Figure 4.**
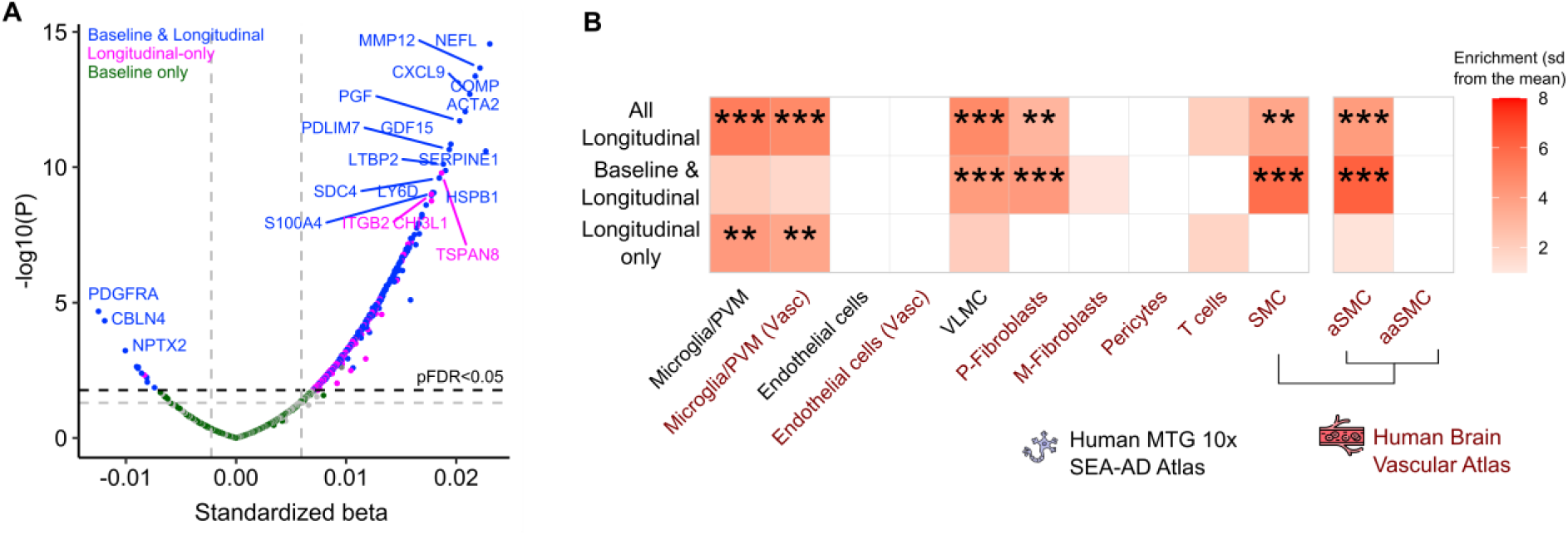
Differential abundance and cell-type enrichment in longitudinal WML progression. **(a)** Volcano plots showing differentially abundant proteins (DAP) associated at baseline with WML progression over time. Linear mixed effect models were adjusted for age, sex, average protein level, and intracranial volume, with a protein*time interaction included in the model. The dashed lines represent p<0.05 (gray) and pFDR<0.05 (black) significance thresholds, and proteins below the pFDR<0.05 threshold were considered significant. Protein color-coding indicates significance relative to WML at baseline: proteins associated with WML both at baseline and longitudinally are highlighted in blue; those exclusively associated with longitudinal WML are in magenta, and proteins associated solely with WML at baseline are shown in dark green. For clarity, only the top upregulated and downregulated proteins from the first two groups are labeled. **(b)** Cell-type enrichment analyses based on single-cell transcriptomics data from the Human MTG 10x SEA-AD and the Human Brain Vascular Atlas for different DAP categories using the Expression Weighted Cell Type Enrichment (EWCE) package. For cell-type enrichment analyses the 1388 Olink proteins were used as background. Not all cell types are shown due to non-significant enrichment. *corresponds to p_FDR_<0.05, **corresponds to p_FDR_<0.01, ***corresponds to p_FDR_<0.001. Abbreviations: OPC = oligodendrocyte precursor cells, PVM = perivascular macrophages, VLMC = vascular leptomeningeal cell, P-Fibroblasts=perivascular fibroblasts, M-Fibroblasts = meningeal fibroblasts, SMC = smooth muscle cell, aSMC = arterial smooth muscle cell, aaSMC=arteriolar smooth muscle cell

### Neuronal and OPC-associated proteins partly mediate the association between WML and cognitive decline

Since WML are associated with cognitive decline, especially in executive functions (Fig. 5a), we next assessed the possible mediation of DAP on the association between WML at baseline and longitudinal cognitive decline. We studied the impact of these proteins on changes in executive function (Trail Making Test-A (TMT-A), Trail Making Test-B (TMT-B) and the Symbol Digit Modalities Test (SDMT), Fig. 5, Supplementary Table 5) and global cognition (the modified Preclinical Alzheimer Cognitive Composite, mPACC, Extended Data Fig. 5). We found that proteins like CSTB and NEFL, which are upregulated in WML, alongside ECM and cell adhesion-related proteins (SMOC2, MMP10, TMSB10), exhibited a significant partial mediation effect on rate of executive function decline, each contributing over 10% to the mediating effect (Fig. 5b). Similarly, key downregulated proteins associated with synaptic function, such as NPTX2, NPTX1, and CBLN4, were major contributors to this mediation. Cell enrichment analysis of proteins that were negatively associated with WML and that mediated cognitive decline showed that these were predominantly neuronal, including glutamatergic and GABAergic subtypes, and OPC-related (Fig. 5c). Proteins that had no mediating effect were primarily observed in OPCs. While proteins positively associated with WML and mediating cognitive decline did not exhibit significant cell-type enrichment, the non-mediating proteins were enriched in VLMC, perivascular fibroblasts, arterial smooth muscle cells and microglial cells. These results were largely reproduced when looking at change in global cognition instead of executive function (Extended Data Fig. 5).

**Figure 5.**
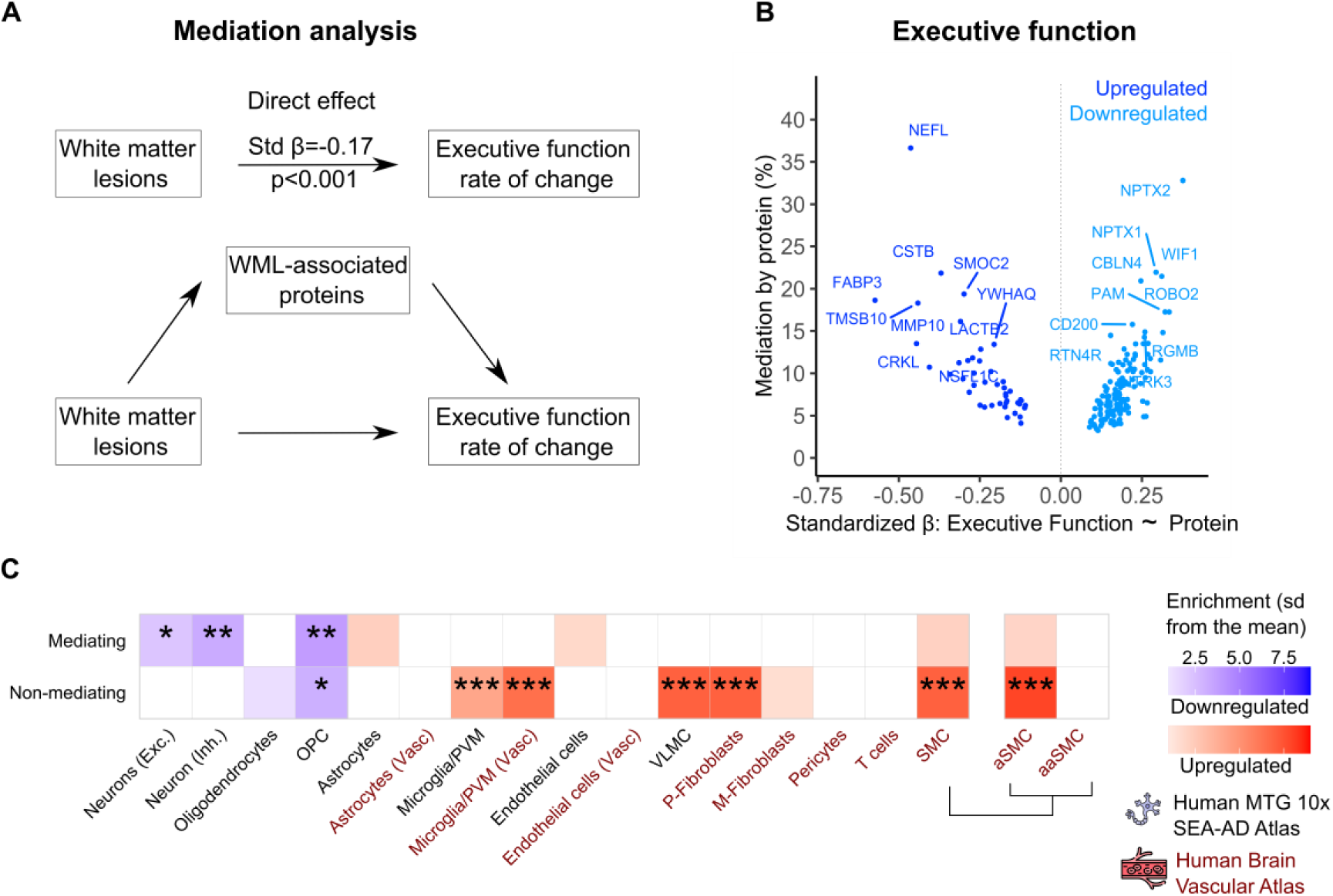
Characterization of WML-associated proteins mediating the relationship between WML and cognitive decline as measured by executive function. **(a)** Path diagram of the mediation analysis models. **(b)** Volcano plots showing the extent to which WML-associated proteins mediate the association between WML and executive function rate of change by the standardized beta values derived from the association between proteins and the executive function rate of change. The top 10 upregulated and top 10 downregulated proteins with the highest mediation effect are labeled. **(c)** Comparison for cell-type enrichment between WML-associated proteins that mediate and those that do not mediate the interaction between WML and executive function rate of change based on single cell transcriptomics data from the Human MTG 10x SEA-AD and the Human Brain Vascular Atlas using the EWCE package. For this analysis, the 1388 Olink proteins were used as background. *corresponds to p_FDR_<0.05, **corresponds to p_FDR_<0.01, ***corresponds to p_FDR_<0.001. Abbreviations: OPC = oligodendrocyte precursor cells, PVM = perivascular macrophages, VLMC = vascular leptomeningeal cell, P-Fibroblasts=perivascular fibroblasts, M-Fibroblasts = meningeal fibroblasts, SMC = smooth muscle cell, aSMC = arterial smooth muscle cell, aaSMC=arteriolar smooth muscle cell

### CSF DAPs are consistent across independent cohorts

To ensure robustness and generalizability of our findings regarding WML-associated DAP, we sought validation in two additional independent cohorts with CSF proteomics data (using WML volume as a continuous variable, Extended Data Table 2, Extended Data Fig. 6). The first included 383 participants from the independent Swedish BioFINDER-1 study, all of whom had both CSF proteomics and MRI data. Our analysis focused on a subset of CSF proteins (N=199) measured with Olink-based technology that overlapped with those examined in our BioFINDER-2 study. The second validation cohort was derived from the Alzheimer’s Disease Neuroimaging Initiative (ADNI), which included 729 participants, and used the aptamer-based SOMAscan 7K (v4.1) platform. Linear regression models revealed a high degree of concordance in the differential protein abundance between the BioFINDER-1 and BioFINDER-2 cohorts, with a 68% agreement (Fig. 6a, Supplementary Table 6). Similarly, a 63% concordance was observed between the ADNI cohort and BioFINDER-2 (Fig. 6b, Supplementary Table 7). The correlation of standardized beta values revealed robust associations, with a strong correlation coefficient of 0.84 between BioFINDER-1 and BioFINDER-2 (Fig. 6c). Additionally, a correlation of 0.65 between the ADNI cohort and BioFINDER-2 (Fig. 6d) indicated a high level of consistency in DAP patterns even in this cross-platform context.

**Figure 6.**
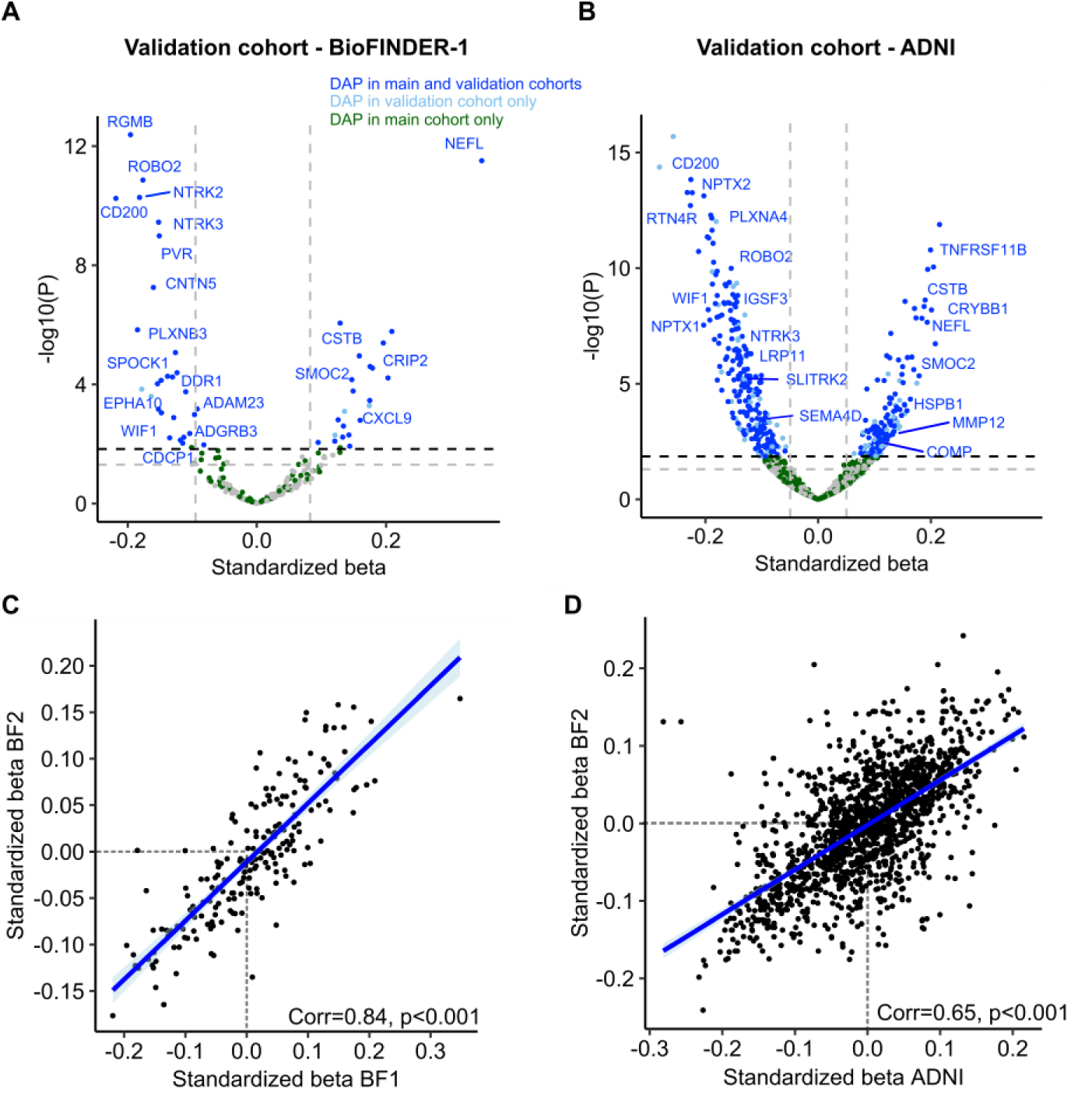
Validation of differentially abundant proteins in CSF in two independent cohorts. (**a,b)** Volcano plots showing differentially abundant proteins (DAP) from the **(a)** BioFINDER-1 validation cohort or **(b)** Alzheimer’s Disease Neuroimaging Initiative (ADNI) validation cohort, analyzed with WML as a continuous variable, adjusting for age, sex, intracranial volume, and average protein level. Color-coding reflects significance across cohorts: proteins significant in both the main and validation cohorts are shown in blue, those significant only in the validation cohort are depicted in pale blue, and proteins significant solely in the main cohort are shown in green. Labels highlight the top 20 differentially abundant proteins in BioFINDER-2. (**c,d)** Scatter plots showing the correlations between standardized beta values from linear regression analysis for proteins overlapping between **(c)** BioFINDER-1 or **(d)** ADNI and BioFINDER-2.

### DAPs in plasma predict future cerebrovascular events and infarcts in a population-based setting

We next investigated whether CSF proteins associated with cSVD outcomes in BioFINDER-2 could be validated in plasma. Plasma proteomic analyses in BioFINDER-2 were conducted using the SOMAscan 7k platform (n=1,599 subjects) and the Olink platform (n=694 subjects). Several proteins were successfully validated in plasma for each cSVD outcome (Supplementary Table 8 and 9). Notably, MMP7, MMP12, TNFRSF11B, GDF15, WFDC2 and NEFL were associated with white matter lesions in plasma (Fig. 7a). Interestingly, some proteins exhibited contrasting trends between CSF and plasma. For instance, S100A4, which had one of the highest standardized beta coefficients for WML in CSF, was notably decreased in plasma. Conversely, WFDC2 showed the opposite pattern, being elevated in plasma but reduced in CSF. Overall, we identified 90 proteins in plasma related to cSVD outcomes in BioFINDER-2, 73 of which were also present in the UK Biobank. This subset, referred to as the “plasma cSVD set”, was further investigated in the UKBB dataset in relation to its extensive data on cerebrovascular outcomes (Fig. 7b).

**Figure 7.**
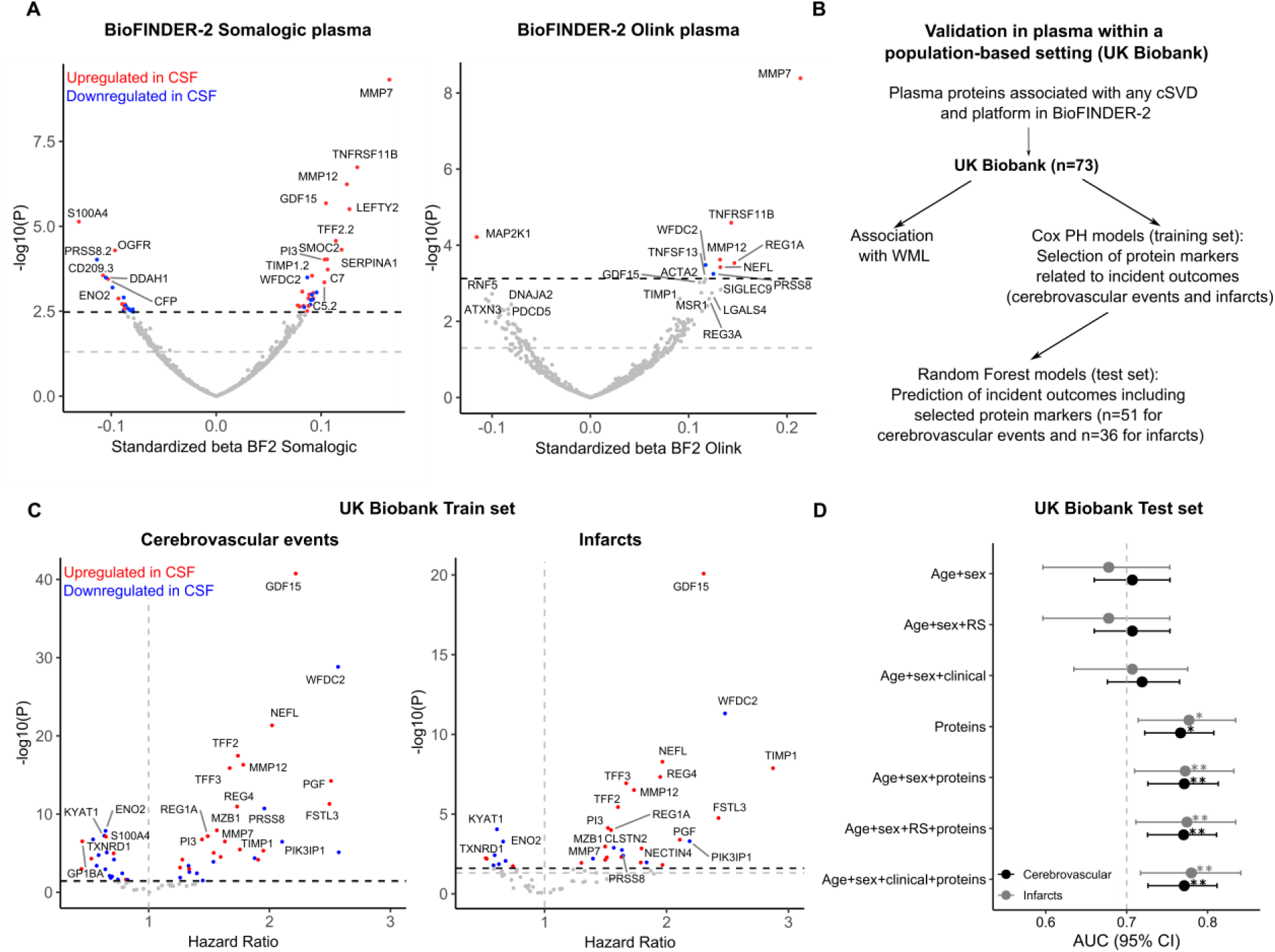
Validation of DAP in plasma and their association with and predictive value for incident cerebrovascular outcomes. **(a)** Volcano plots showing DAP in plasma from the BioFINDER-2 cohort, assessed using Somalogic or Olink platform, analyzed with WML as a continuous variable, adjusting for age, sex, intracranial volume and average protein level. Labels highlight the top 20 DAP. Significant proteins between the two analyses for all cSVD were retained for the UK Biobank analysis. **(b)** Workflow for analysis in UK Biobank. **(c)** Volcano plots illustrating the prospective associations between the plasma cSVD set and the risk of cerebrovascular events or infarcts. The top 20 proteins are highlighted, with color coding indicating whether they were upregulated (red) or downregulated (blue) in CSF. **(d)** Forest plot showing the area under the curve (AUC) values with 95% confidence intervals (CI) for models predicting cerebrovascular events and infarcts. The models were validated using a Random Forest classifier on the test dataset. Different combinations of predictive features are presented, with a model incorporating age, sex and risk score as the base model. Outcomes (cerebrovascular events vs. infarcts) are differentiated by point and line color. AUC for the protein-based and combined models were compared to the baseline model using DeLong’s test. RS = Risk Score. *corresponds to ROC p_FDR_<0.05, **corresponds to ROC p_FDR_<0.01.

We first examined the relationship of the 73-protein plasma set with WML, followed by an evaluation of whether these proteins were associated with the risk of cerebrovascular events or infarcts. Several key proteins from the plasma cSVD set, including GDF15, NEFL and PGF, were associated with WML in the UK Biobank (pFDR<0.05, Extended Data Fig. 7). MMP12, MMP7 and TNFRSF11B showed nominal significance (p<0.05) but did not pass FDR correction.

Next, we divided the UK Biobank dataset into a training set (80%) for inferential analysis, and a left-out test set (20%) to use for prediction analysis (Extended Data Table 3). In the training set, we used Cox proportional hazard ratios to identify the proteins in plasma associated with the risk of incident cerebrovascular outcomes over a 5-year period. 51 of the 73 proteins were significantly associated with cerebrovascular events, and 36 with infarcts (Fig. 7c). Among these, GDF15 emerged as a top hit in both analyses, with WFDC2, NEFL, PGF and MMP12 also identified as key proteins in both analyses. TIMP1 exhibited the highest hazard ratio for infarcts, while its association with any cerebrovascular events was comparatively weaker. We then evaluated if the combinations of those proteins (51 for cerebrovascular events and 36 for infarcts) could further improve the prediction of both outcomes in the test set using a random forest (Fig. 7d). We compared models with various feature combinations, including protein-only and protein-inclusive models, against a base model comprising age, sex and a clinical risk score, the latter serving as benchmark in clinical practice (i.e., the American Heart Association Stroke Risk Assessment Score). Additionally, we evaluated models using the clinical variables that comprise the risk score, analyzed as continuous or binary variables. Across all feature sets, the addition of proteins significantly improved predictive performance beyond the base model (DeLong test, p<0.05). For cerebrovascular events, the base model had an AUC of 0.72 (95% CI: 0.68–0.77), which improved to 0.77 (0.72–0.81) with the addition of proteins alone and remained 0.77 (0.72–0.81) when age, sex, and clinical covariates were added alongside proteins. For infarcts, the base model had an AUC of 0.68 (95% CI: 0.60–0.75), increasing to 0.78 (0.71–0.83) with proteins alone, and reaching 0.78 (0.72–0.84) with the addition of age, sex, and clinical covariates. Several proteins, such as GDF15 and MMP12, demonstrated high SHAP values, indicating their strong contribution to the model’s predictive performance for cerebrovascular events and infarcts (Supplementary Fig. 12). Notably, in the model combining age, sex, clinical variables and proteins, only protein features were included in the top 15 most important features for either model (Extended Data Fig. 8).

## DISCUSSION

This study thoroughly investigates CSF proteomic alterations in various cSVD manifestations in several large cohorts, while also drawing parallels with broader cerebrovascular disease, including large artery disease. Earlier work focused primarily on isolated cSVD manifestations, often centered on WML, with limited protein panels assessed mostly in blood. These studies identified significant associations between cSVD pathology and endothelial dysfunction (ICAM-1^19^, E-selectin^20^, PGF^21,22^), inflammation (IL-6^23^), clotting pathways (vWF, CSTB, LTBR, TNFRSF11B)^24^ and alteration of neuronal structural integrity (NEFL^25^), which were validated in our study. We identify novel candidate biomarkers and provide an enriched expression profile that deepens the understanding of the pathophysiological framework of cSVD. We have uncovered both unique and shared patterns across cSVD conditions, with e.g. MMP12 emerging as an early and robust indicator. Our findings also highlight a critical role for arterial smooth muscle cells and shed light on the dynamics between disrupted metabolic, vascular, immune, and neuronal mechanisms in cSVD. Furthermore, several key proteins were validated in plasma, with a subset improving 5-year risk stratification for cerebrovascular events and infarcts above and beyond current clinical benchmarks. In summary, we have identified proteins, and biological processes present in cSVD that hold promise as novel therapeutic targets and biomarkers for these diseases.

Several proteins linked to extracellular matrix (ECM) organization and smooth muscle cell function were elevated across CVD manifestations. In cSVD, in addition to ECM-related proteins such as MMP12 and ELN (further detailed below), matricellular proteins like POSTN and COMP were also altered. COMP, an ECM glycoprotein essential for collagen assembly and ECM stability, and has been linked to atherosclerosis, plaque area and vulnerability, and may induce tissue fibrosis^26^, while POSTN, upregulated in vascular injury, promotes ECM remodeling and inflammation^27,28^. In addition, the upregulation of PDLIM7 and ADAM8 suggests shared mechanisms in cSVD, involving ECM remodeling, vascular repair, and altered cell-matrix communication. These findings point to a common cluster of upregulated proteins driving vascular integrity and remodeling in cSVD pathophysiology.

Among these shared proteins, MMP12 seems to be a key early player in the onset of cSVD, marked by an early and substantial increase, and association with WML progression. While previous research has shown associations between MMP2, MMP3 and MMP9 and vascular cognitive impairment^29,30^, we did not find an association with MMP3, and technical constraints prevented the evaluation of MMP2 (not present in the Olink panel) and MMP9 (above LOD in <70% of subjects). Interestingly, we found an association of other MMPs with WML, such as MMP1, MMP7, MMP8 and MMP10. MMPs can influence cSVD by proteolyzing cerebrovascular basement membranes, altering tight junctions, and activating bioactive molecules like clotting factors and signaling proteins^31^. MMP12 plays a key role in breaking down elastin and other ECM components^32^, which in pathological conditions may lead to compromised vessel wall integrity and elasticity. Elevated MMP12 within the small arteries of subjects with cSVD further suggests its involvement in vessel wall remodeling and arterial stiffening, as supported by evidence from human aortic and animal studies^33^. This aligns with our observations of increased ELN levels across cSVD pathologies, suggesting the breakdown of elastic fibers and elevated levels of various collagen types, especially with WML. Arterial stiffness can increase endothelial permeability, promote macrophage adhesion, and can lead to smooth muscle proliferation and vessel remodeling^34^. Additionally, MMP12 also modulates neuroinflammation through its effects on macrophages and neutrophils^35,36^, suggesting that its upregulation is linked to early vascular injury responses and subsequent pathogenic cascades.

MMP12 was co-abundant within a network of proteins that were elevated in cSVD (Module 5), whose levels increased during the early stages of WML and microbleed pathology. This highlights the critical role of endothelial and smooth muscle cell dysfunction at the onset of cSVD pathogenesis, with proteins likely involved in vascular remodeling. Such remodeling might include a shift toward a synthetic phenotype with reduced contractility and increased proliferation, as observed in animal models of hypertension^37^, and/or gradual degeneration and loss of smooth muscle cells^38^. The presence of cSVD-associated proteins like HSPB1, HSPB6, MMP12, PDLIM7, SERPINE1, and CSTB within this module suggests that oxidative stress, endothelial dysfunction, and vascular remodeling may collectively drive cSVD development and progression. Additionally, a strong association with Module 10, which includes proteins like SMOC2^39,40^ (involved in ECM degradation and smooth muscle cell migration), and VWF^41,42^ (a marker of endothelial dysfunction), points to a combined influence of metabolic and vascular dysfunction in promoting vascular damage and facilitating the entry of potentially harmful toxins and immune cells into the brain^43^.

The link between inflammation and vascular damage is well established, though the direction of causality remains debated^44^. WML were uniquely associated with VLMC- and perivascular fibroblast-enriched proteins, suggesting their role in immune cell recruitment, leukocyte migration, and local inflammation through chemokine signaling and extracellular matrix production^45,46^. Our findings highlight immune cell involvement in WML progression, with strong associations observed for microglia- and perivascular macrophage-associated proteins, such as ITGAM, ITGB2, CHI3L1 and IL15. This suggests a strong pro-inflammatory response, which might accelerate cell death, and the development and expansion of WML. Indeed, increased levels of serum CHI3L1 have recently been associated with cSVD and with the destruction of white matter macroscopic and microstructure, further associated with cognitive deficits^47^. In multiple sclerosis, CHI3L1 was associated with neuronal deterioration, affirming its contribution to the progression of the disease^48^. In a rodent model, microglial inhibition via minocycline reduced white matter damage^49^, and its effects on microglia activation and BBB permeability in cSVD patients are currently under investigation in a phase-2 clinical trial^50^. Future research should explore microglial states and stage-specific contributions to cSVD.

This study observed widespread downregulation related to neuron- and OPC-related proteins in WML, to a lesser extent in deep microbleeds and subcortical infarcts/lacunes, and strikingly lower levels in lobar microbleeds and cortical/cerebellar infarcts. This suggests that proteomic response to cerebrovascular injury may be highly localized, with subcortical regions and white matter more susceptible to disruptions and leading to a broader proteomic downregulation. Furthermore, WML-associated protein downregulation appears to have different impacts on cognitive outcomes. Specifically, a subset of proteins abundant in OPCs that were not linked to cognitive decline may be involved in the myelination process and serve broader roles including proliferation, migration and differentiation of OPCs (ERBB3/ERBB4)^51^ and extracellular matrix organization (TIMP4)^52^. While these proteins are vital for maintaining efficient neuronal signaling, their direct impact on cognitive functions may not be immediate, and the brain may partially compensate for reductions in myelination efficiency.

Conversely, downregulated neuronal proteins, and some OPC-enriched proteins, are more directly linked to cognitive decline due to their role in synaptic function and plasticity. Key mediators include CBLN4 (synaptic organization and plasticity^53^), and NPTX1/NPTX2 (synaptic activity biomarkers and potential CSF biomarkers for synaptic dysfunction in neurodegeneration^54^). Additionally, EphA10 and EphB6, both pseudokinases of the Eph receptor family, are emerging but underexplored mediators of cognitive decline. While the Eph family is recognized for its involvement in synaptogenesis, synaptic plasticity and contribution to neurological diseases characterized by memory impairments^55^, EphA10 and EphB6 are not well characterized. Recent studies suggest EphB6 is present in both excitatory and inhibitory synapses^56^, highlighting its potential role in synaptic communication and the need for further investigation.

Several proteins *upregulated* in WML and mediating WML-associated cognitive decline also show elevated levels in an AD context^57,58^, but remain associated with WML when adjusting for AD. Notable mediators include CSTB, a lysosomal cysteine protease implicated in neurodegeneration^59^ and NEFL, a key cytoskeletal component reflecting neuronal damage and associated with neurodegenerative diseases^60^. Other proteins such as SMOC2, MMP10 and TMSB10, associated with cognitive decline, highlight the role of ECM modulation and cell adhesion^39,61,62^ in cognitive function.

Given that the distinct topography of microbleeds is considered to reflect their underlying etiology, we revealed a compelling separation of molecular pathways. Deep microbleeds, commonly associated with hypertensive arteriosclerosis^63^, share a proteomic profile with other cSVD manifestations, including WML and subcortical infarcts/lacunes, with key proteins like MMP12, TNFRSF11B and SERPINE1 implicated in arterial stiffness and atherosclerosis pathogenesis^64–67^. In contrast, lobar microbleeds, linked to cerebral amyloid angiopathy (CAA), exhibit a distinct profile, with protein like ITGB2 and GLOD4 associated with amyloid pathology^68^. Nevertheless, proteins shared between lobar microbleeds and WML, such as CHIT1, CCN1 and MMP10, suggest overlapping pathways of vascular injury and tissue repair. These findings support a model in which deep microbleeds align with cSVD pathways, while lobar microbleeds parallel more closely amyloid pathways.

In the context of microbleed pathology, CHIT1 has emerged as a key protein, showing early and progressive increases as the pathology worsens. As a promising novel marker of acute or chronic inflammation, its expression has been mostly ascribed to microglia and infiltrating macrophages^69^. CHIT1 is elevated in CSF in both AD^70,71^ and cerebrovascular dementia^72^, but its association with microbleeds persisting after adjusting for AD pathology. Interestingly, in multiple sclerosis, baseline CSF CHIT1 levels reflect microglial activation early in the disease and correlated with subsequent disease progression^73^. Its early increase in microbleeds suggest CHIT1 may serve as a marker as an early marker in the inflammatory response in microbleeds, warranting further investigation into its role in immune response and cSVD.

In cortical and cerebellar infarcts, which are indicative of large artery disease, RhoC emerged as a prominent marker. As a member of the Rho GTPase subfamily, RhoC regulates vascular homeostasis by modulating endothelial cell migration, proliferation, and permeability^74,75^, but remains unexplored in this context compared to RhoA and RhoB, which are upregulated in focal cerebral infarctions from as early as 2 days to as long as 38 months post-infarction^76^. In infarcts, particularly subcortical infarcts and lacunes, downregulation of astrocyte-enriched proteins, including RGMA, LRP1 and MEGF10, suggest altered astrocyte functions. These proteins are linked to reactive astrogliosis, scar formation after stroke^77^, and phagocytosis^78,79^, with LRP1 loss shown to mitigate demyelination and reverse white matter lesions.

After establishing key protein associations in CSF, we next focused on plasma proteomics to investigate whether similar patterns can be observed. We validated several proteins in plasma, with MMP7 emerging as top hit, showing strong associations with white matter lesions. Like MMP12, MMP7 plays a pivotal role in ECM degradation but is also critical for maintaining blood-brain (BBB) integrity^80^. Elevated MMP7 levels have been observed in conditions with BBB compromise, such as multiple sclerosis (MS) and traumatic brain injury (TBI), where its serum levels correlate with MRI-detected BBB dysfunction^81,82^. Given its role in ECM remodeling and BBB integrity, MMP7 has been proposed as a potential marker for BBB integrity, further underscoring its relevance in cerebrovascular disease.

To focus on the most robust and clinically relevant associations, we evaluated the predictive value of plasma proteins validated first in CSF, for cerebrovascular outcomes in UK Biobank. This cross-fluid approach revealed a subset of plasma proteins that were associated with the risk of cerebrovascular events and significantly improved 5-year risk stratification, surpassing models based on age, sex, and stroke risk score. Among the most prominent proteins were GDF15, WFDC2, NEFL and MMPs outlined previously. These findings align with a recent study demonstrating the utility of a protein-based risk score for 10-year infarct prediction^83^ but go further by linking specific proteins to cSVD pathology and associating them with incident cerebrovascular outcomes. This highlights the predictive value of proteomic markers and underscores their potential to improve early risk identification and support personalized prevention strategies for cerebrovascular events and infarcts. Further research is needed to validate these findings across diverse populations and to explore the integration of proteomic biomarkers into clinical practice.

The cohort’s recruitment from southern Sweden, primarily composed of individuals of European descent narrows ethnic and racial representation. This points to the need for inclusion of more diverse populations in future studies, especially considering recent evidence of ancestry-specific protein co-abundance modules^84^ and findings that cSVD affects certain populations disproportionately^85^. The validation of many DAP in separate datasets, the Swedish BioFINDER-1 study, the UK Biobank cohort and the American ADNI study—with ADNI employing the SOMAscan proteomics platform—demonstrates the robustness of our findings despite varied methodologies. The concordance between the SOMAscan and Olink platforms supports the credibility of these protein signatures as biomarkers for cSVD, suggesting their applicability across diverse patient groups and detection technologies. Furthermore, to gain further insight into the relationship between CSF markers and cSVD progression, it is imperative to pursue longitudinal studies that track protein level changes over time.

In conclusion, using a multi-omics approach, we showed that cSVD manifestations present both shared and distinct proteomic signatures, revealing diverse pathophysiological profiles. These findings compel a reevaluation of the traditional view of ‘cerebral small vessel disease’ as an umbrella term towards a more nuanced classification into distinct ‘cerebral small vessel diseases’, each characterized by unique biomarker profiles. We have highlighted a critical role for arterial smooth muscle cells in cSVD pathophysiology and identified candidate biomarkers in CSF and plasma, which hold promise for novel diagnostic and risk stratification applications. Furthermore, the identification of key molecular pathways and early biomarkers highlights opportunities for targeted therapeutic development, supporting efforts to improve the clinical management of cSVD.

## METHODS

### Participants – BioFINDER-2 cohort

We included participants from the ongoing, prospective Swedish BioFINDER-2 cohort (NCT03174938). All participants were recruited from the south of Sweden at Skåne University Hospital or the Hospital of Ängelholm, Sweden, and the data were acquired between May 2017 and December 2023. The main inclusion and exclusion criteria has been described in detail elsewhere^86^. All participants included in this study had complete CSF proteomic data and MRI measurements available at baseline, with the final dataset including 1670 subjects. The patients ranged from adults with intact cognition or subjective cognitive decline (both included in the CU group), mild cognitive impairment (MCI), and dementia. Causes of cognitive impairment included AD and non-AD neurodegenerative diseases. The group of non-AD neurodegenerative diseases included Parkinson’s disease (PD) and vascular dementia (VaD) patients. The underlying etiology was established based on primary criteria for each condition, either at baseline or during a follow-up visit. Written informed consent was obtained from all participants prior to entering the study. Ethical approval was obtained from the Regional Ethical Committee in Lund, Sweden.

### MRI as measure of cerebrovascular pathology

MRI was performed using a 3 Tesla Siemens MAGNETOM Prisma scanner, equipped with a 64-channel receiver coil array (Siemens Healthcare, Erlangen, Germany). Imaging sequences included a whole-brain T1-weighted anatomical magnetization-prepared rapid gradient echo sequence (MPRAGE; TR = 1900 ms, TE = 2.54 ms, voxel size = 1 × 1 × 1 mm3), a T2-weighted fluid-attenuated inversion recovery sequence (FLAIR; TR = 5000 ms, TE = 393 ms, matching resolution with the T1-weighted image), and a 3D, multi-gradient-echo pulse sequence (TR = 24 ms; TEs = 5.00, 8.80, 12.60, 16.40, and 20.20 ms with monopolar/fly-back readout gradients, flip angle = 15°; voxel size = 0.8 × 0.8 × 1.4 mm3).

Following established criteria, microbleeds and infarcts were identified and counted via visual inspection by an experienced neuroradiologist (D.W.), from the T2* and the FLAIR images, respectively^5,87^. We categorized microbleeds and infarcts as either 0 (absent) and ≥1 (present). For certain posthoc analysis, microbleed counts were treated as a continuous variable and log-transformed for analysis. Microbleeds were further classified based on their localization into lobar and deep, while infarcts were categorized as either cortical/cerebellar or subcortical infarcts/lacunes. White matter lesions (WML) were segmented using the longitudinal Sequence Adaptive Multimodal SEGmentation (SAMSEG) tool from FreeSurfer (version 7.1) using the T1 and FLAIR images as input and quantified^88,89^. When treating WML volume as a continuous variable, we applied min-max scaling to rescale the range to fall between 0 and 1. In the main analysis, WML was treated as a binary variable to maintain uniformity in comparison to the other cSVD. Based on the histogram, we divided the WML data into tertiles, such that the top tertile effectively captured WML-positive individuals (Supplementary Fig. 1). The cutoff for this top tertile was 0.497% of ICV which is consistent with previous studies using a 0.5% of ICV cutoff^90,91^. Baseline data on microbleeds were missing for 51 participants. MRI follow-ups were conducted 1, 2, 4 and 6 years after the initial baseline measurement, for an average of 2.94±1.02 years of follow-up.

### CSF proteomic measures

CSF samples were collected during baseline visits and were subjected to analysis using the Olink proteomic assay, a highly sensitive and specific antibody-based Proximity Extension Assay (PEA^TM^ technology)^92^. This method involves the use of multiplexed oligonucleotide labeled antibody probe pairs that bind to their respective target protein. Protein expression levels were reported as normalized protein expression (NPX) values on a log2 scale, providing a relative quantification unit. We used the Olink Explore 3072 panel, allowing for quantification of 2943 proteins within eight multiplex protein panels, each containing 367-369 proteins (Oncology I and II, Neurology I and II, Cardiometabolic I and II, and Inflammation I and II). To ensure rigorous quality control (QC) in line with Olink technology standards, samples were randomly distributed across plates, and each plate included relevant controls. Samples failing to meet stringent manufacture-provided QC criteria were labeled with a warning. A lower limit of detection (LOD) was defined as three standard deviations above the control-based background for each assay. Our analysis was focused on 1388 proteins demonstrating strong expression, with levels exceeding the limit of detection (LOD) in ≥70% of the participants^68^. Protein values with Assay or QC warnings, as well as outliers defined as values ±5 standard deviations from the mean, were excluded from the analysis. For analysis, we retained all data points below the LOD, following Olink recommendations^16,93^. Building on our prior study that identified the existence of inter-individual variability in average CSF protein levels^94^, we included a measure of average overall protein level as a covariate in our analysis. This was computed as the mean z-scored NPX value for proteins with high detection rates (n=1199, including proteins exceeding the LOD in over 90% of participants).

### Cell-type and functional enrichment analysis

For cell-type enrichment, we used two sources with single-nucleus RNA-sequencing data: 1) single-nucleus transcriptomes from 166,868 nuclei in the middle temporal gyrus (MTG) obtained from the SEA-AD (Human MTG 10x SEA-AD 2022, https://portal.brain-map.org/atlases-and-data/rnaseq/human-mtg-10x_sea-ad), from five post-mortem human brain samples and 2) the Human Brain Vascular Atlas^95^, which profiles major vascular and perivascular cell types of the human brain using 143,793 single-nucleus transcriptomes from the hippocampus and cortex from eight post-mortem samples. We acquired the Seurat objects for both atlases and used the function AverageExpression (R package Seurat version 4.3.0) to determine cell type expression levels. By leveraging the class and subclass annotations from the Human MTG 10x SEA-AD data, we computed the average expression from both non-neuronal cells (microglia, astrocytes, oligodendrocytes, oligodendrocyte precursor cells, vascular leptomeningeal cells, and endothelial cells) and neuronal cells (GABAergic neurons and glutamatergic neurons), after removing the annotations labeled as “None”. Following the same procedure, we calculated the average expression in major vascular and perivascular cell types from the Human Brain Vascular Atlas, including endothelial cells, smooth muscle cells (distinguishing between arterial and arteriolar smooth muscle cells in a subsequent analysis), pericytes, astrocytes, macrophages, T cells, perivascular and meningeal fibroblasts. The percentage expression across the cell types was then calculated based on their average expression levels. For assessing cell-type enrichment statistically, we used the EWCE^96^ (Expression Weighted Cell Type Enrichment) package in R (v1.6.0) to determine whether a specific set of proteins (DAP or modules) showed significant enrichment within certain cell types beyond what would be expected by chance. This was done against a background of the 1388 Olink proteins, using 10,000 bootstrap iterations for analysis.

Functional enrichment analysis on relevant set of proteins (DAP and modules) was performed using WebGestalt^97^, with the 1388 Olink proteins as background set. We performed Over-Representation Analysis (ORA) with ontology sources from Gene Ontology (GO) Biological Processes (BP), Cellular Component (CC) and Molecular Function (MF). We report terms with pFDR<0.05 (Benjamini–Hochberg method), after having applied redundance reduction of the enrichment categories. All pFDR<0.05 terms are reported in the Supplementary table.

### Co-abundance modules analysis

To identify modules of co-expressed proteins, we employed the SpeakEasy2 consensus clustering algorithm, which simultaneously applies both top-down and bottom-up approaches to detect robust protein communities^18^. We utilized previously developed SpeakEasy clusters derived from 1,331 proteins^98^ and integrated additional proteins by correlating their expression values with cluster centroids, assigning them to the closest matching cluster on a winner-takes-all basis.

### Immunofluorescent staining from postmortem brain tissue

Immunofluorescent staining of frontal lobe white matter tissue was performed on a cohort of six SVD cases (n=6, age 81-90+ years; three women, severe frontal white matter rarefaction (e.g. grade 3) and at least one frontal lobe microscopic cortical infarct, Braak II-IV) and six non-SVD controls (n=6, age 85-90+ years; three women, low grade frontal white matter rarefaction (e.g. grade 0-1), no old microscopic or lacunar infarcts, Braak II-IV) from the Arizona Study of Aging and Neurodegenerative Disorders and Brain and Body Donation Program at Banner Sun Health Research Institute^99^. The cases were evaluated according to CWMR grading system^100^ and National Institute on Aging and Reagan Institute (NIA-RI) criteria^101^, based on NFT Braak stages (I-VI)^102^ and Consortium to Establish a Registry for Alzheimer disease (CERAD) neuritic Aβ-plaque scores^103^.

Informed consent for the use of brain tissue, plasma, and clinical data for research purposes was obtained from all subjects or their legal representatives in accordance with the International Declaration of Helsinki^104^. Procedures for the collection of brain tissue have been described earlier ^86,105^. The tissue collection protocols were approved by the Western Institutional Review Board of Puyallup, Washington (USA) and the Swedish Ethical Review Authority approved the study. Directly after autopsy, the brain tissues were fixed in 10% neutral buffered formalin solution containing 4% formaldehyde, followed by 48h in 2% dimethyl sulfoxide/10% glycerol and another 48h in 2% dimethyl sulfoxide/20% glycerol. The tissue was then sectioned in 40μm free-floating sections and stored in cryoprotectant at 4°C until used for immunostaining.

To assess MMP12 localization by immunofluorescence, sections from control and SVD cases were stained with antibodies directed against MMP12 and laminin. Sections were incubated in blocking solution (5% goat serum, 0.25% Triton X in KPBS) for 1 hour, followed by mouse anti-laminin (1:200, Dako) and rabbit anti-MMP12 (1:500, Biorbyt) in blocking solution for 48 hours at 4°C. After washing, sections were incubated with secondary antibodies (1:500, Dylight 549 goat anti-mouse and 1:500, Alexa 488 goat anti-rabbit, Thermo Fisher Scientific) for 2 hours at room temperature. All sections were treated with Sudan Black (1% in 70% ethanol, Sigma-Aldrich) for 5 minutes, rinsed, and mounted with Vectashield containing DAPI (Vector Laboratories).

To analyze MMP12 expression within arteries, images of five white matter arteries (outer diameter of 150–400 μm) were captured using a 20x objective (Olympus BX41). The area fraction of MMP12-stained regions within laminin-stained regions was subsequently quantified using Fiji/ImageJ software. The laminin-stained area was manually delineated as the region of interest, and thresholds for each staining channel were adjusted manually based on the intensity distribution to ensure accurate separation of MMP12 and laminin signals. The area fraction of MMP12 staining within the laminin-stained region was calculated by dividing the thresholded MMP12-positive area by the total laminin-positive area. To minimize bias, the observer conducting the thresholding and measurements was blinded to the sample identities throughout the analysis. Group differences were analyzed using Mann-Whitney U test.

### Validation of key DAP in CSF in independent datasets

Two separate cohorts were used to validate key proteins identified in CSF in the BioFINDER-2 cohort: BioFINDER-1 and Alzheimer’s disease Neuroimaging Initiative (ADNI). The BioFINDER-1 study (registered under NCT01208675) is comprised of cognitively unimpaired (CU) individuals and individuals with mild cognitive impairment (MCI). The methodologies for participant selection and the criteria for inclusion or exclusion have been described previously^106^. In BioFINDER-1, the proteomic analysis was conducted on a limited set of proteins from different panels, including Neuro-exploratory, Neurology, Inflammation, and Cardiovascular-III, resulting in an overlap of 199 proteins with the main BioFINDER-2 cohort. These overlapping proteins were used for further analysis. From this cohort, we selected participants who had both proteomic data and MRI data, specifically those for whom data on white matter lesion volumes were available, segmented using SAMSEG. This subset included 383 subjects.

To validate key proteins externally, we used the dataset from Alzheimer’s disease Neuroimaging Initiative (ADNI), an ongoing, multisite observation study aimed at understanding the progression of Alzheimer’s disease (https://adni.loni.usc.edu/). We selected participants from ADNI1, ADNI-GO and ADNI2 who had proteomic and MRI data available, including information on white matter lesions, resulting in a total of 729 individuals. Using SomaScan 7K (v4.1), an aptamer-based proteomics technology, protein levels of ∼7000 proteins were measured, and analyses were performed on 1467 overlapping aptamers with BioFINDER-2. White matter lesion volume was taken from the “ST128SV” variable, which was quantified using FreeSurfer from T1-weighted images. The total intracranial volume was taken from the variable “ST10CV”.

### Validation of key DAP in plasma and predictive modeling in UK Biobank

The next objective was to validate key proteins in plasma within a population-based setting, assessing their relevance to real-world applications. To achieve this, we first identified proteins that translate to plasma using data from the BioFINDER-2 dataset and further evaluated the predictive utility of these proteins for clinically relevant outcomes in the UK Biobank (application number 105777)^107^. First, proteomic analysis in plasma was conducted on BioFINDER-2 subjects using the SOMAscan7k platform (n = 1,599) and/or the Olink Explore HT platform (n = 694), with >95% participant overlap between the CSF and plasma datasets. Quality control and preprocessing were conducted using the same protocols as the Olink dataset, with additional log2 normalization applied to proteins from the SOMAscan platform. After preprocessing, for each model (WML continuous and binary, deep and lobar microbleeds, subcortical infarcts/lacunes and cortical/cerebellar infarcts) we isolated proteins that were: i) significant in the CSF model; ii) additionally present in the plasma database after quality control. To validate DAP from CSF in plasma for each condition, we applied linear models for each cSVD manifestation independently, adjusting for age, sex and average protein levels. Validation analysis was performed separately for Olink and SOMAscan. Given the larger dataset available for SOMAscan and the demonstrated modest concordance between the two platforms^108,109^, we utilized both to maximize the detection of relevant biomarkers and avoid missing potential associations.

Proteins that remained significantly associated with cSVD pathology in plasma across proteomic platforms in BioFINDER-2 were further analyzed in the UK Biobank. The UK Biobank is a large-scale, multicenter prospective cohort study involving approximately 500,000 participants aged 40–69, recruited between 2006 and 2010. Detailed information about the study’s methodology and data collection is available in the UKBB online protocol (www.ukbiobank.ac.uk). In this cohort, 2,922 plasma proteins were measured using the Olink Explore 3072 assay. Following the same protein retention criteria and protein matching, 73 proteins were significantly related to cSVD in plasma in BioFINDER-2 and were included for further analysis in UK Biobank. We will refer to this set of proteins henceforth as the “plasma cSVD set”.

We first examined the association between the plasma cSVD set and WML in UK Biobank, since WML was the only available cSVD marker in the UK Biobank. The analysis included 5,712 participants with both Olink proteomics data and WML volume information derived from MRI scans. WML volumes were obtained from the variable "Total volume of white matter hyperintensities from T1 and T2 FLAIR images," with lesion segmentation performed using the BIANCA tool^110^. Total intracranial volume (ICV) was derived from the variable "Volume of Estimated Total IntraCranial (whole brain)."

As a next step, we aimed to pinpoint proteins from the plasma cSVD set that are associated with incident outcomes in the UK Biobank: infarcts and cerebrovascular events. For this analysis, we utilized a more comprehensive dataset from the UK Biobank, incorporating subjects with both proteomic data and diagnostic information. We split this dataset into a training (80%) and test (20%) subset to enable robust model development and validation (see Extended Data Table 3 for demographic information). In the training set only, Cox Proportional Hazard ratios models were used to preselect relevant proteins associated with two incident outcomes in the UK Biobank, infarcts and cerebrovascular events, occurring within five years. In this manner, the training set was used for feature selection to prioritize proteins for inclusion in predictive models, while the test set was reserved for evaluating machine learning performance. The analysis was conducted using the survival package (version 3.4-0). Models were adjusted for age, sex, and mean protein values.

Infarcts were defined using ICD-10 code I63 (cerebral infarct), while cerebrovascular events were identified using ICD-10 codes I60-I68, which include conditions such as hemorrhages, infarcts, arterial occlusions and stenosis, cerebral amyloid angiopathy (CAA), and other cerebrovascular diseases. These codes were obtained from a UK Biobank field (data field 41202) summarizing distinct primary or main diagnosis codes recorded in participants’ hospital inpatient records. Patients with a history of cerebrovascular events prior to their first visit were excluded from the analysis.

After identifying proteins significantly associated with cerebrovascular events and infarcts, we investigated whether incorporating these proteins could improve the predictive performance of an established stroke risk score in the test set. The covariates used for this analysis were based on the Stroke Risk Assessment Score^111^ from the American Stroke Association. These included blood pressure (average of two readings), fasting glucose levels, body mass index (BMI), total cholesterol, prior diagnoses of atrial fibrillation and diabetes mellitus, personal or family history of stroke, transient ischemic attack (TIA), or heart attack, and smoking status. In accordance with the Stroke Risk Assessment, a composite risk score was calculated by assigning one point for each risk factor present: blood pressure>120/80 mmHg, glucose>100 mg/dL, BMI>25 kg/m², cholesterol>160 mg/dL, atrial fibrillation, diabetes, family or personal history of cerebrovascular events, and smoking, with a maximum score of 8. Missing data in covariates and protein measurements were addressed through systematic imputation: continuous variables were imputed using k-nearest neighbors, and categorical/binary variables were imputed using median values.

Classification accuracy was tested using a Random Forest classifier. Hyperparameter tuning (max depth: [3, 5, 7, 10, 15, 20, 40, 50, None]) was performed on the training set with grid search and five-fold cross-validation, with balanced accuracy as the optimization metric. The final model’s performance on the test set was measured using the area under the receiver operating characteristic curve (AUC). Various feature combinations were tested to predict cerebrovascular events or infarcts within five years. Proteins were modeled alone or alongside the stroke risk score or individual clinical variables (systolic and diastolic blood pressure, glucose levels, MBI, cholesterol, atrial fibrillation, diabetes, family or personal history of cerebrovascular events and smoking) to evaluate their added value in enhancing risk stratification and improving predictive accuracy. Statistical significance of the improvement in model performance was evaluated using DeLong’s test, comparing models to the base model containing age, sex and the stroke risk score. To assess feature contributions, Shapley Additive Explanations (SHAP) values were calculated for the model incorporating age, sex, clinical variables, proteins, and average protein level.

### Statistical analysis

All analyses were performed in R version 4.2.1, Python version 3.9.2 or Graph Pad Prism software v10.3.0. Linear models were used to assess DAP for each cSVD manifestation independently (WML+ vs WML-, Microbleeds+ vs Microbleeds-, Infarcts+ vs Infarcts-), including age, sex and average overall protein levels as covariates. Further analysis distinguished between lobar microbleeds (Lobar microbleeds+ vs Lobar microbleeds-) and deep microbleeds (Deep microbleeds+ vs Deep microbleeds-). Subsequently, significant DAP for each pathology were analyzed in a linear model including all cSVD as independent variables, to determine if the association persisted after adjusting for other cSVD. Only proteins that retained significance for each disease independently were considered in subsequent analyses, including cell-type and functional enrichment. In specified analyses, models were adjusted for presence of clinical AD and PD diagnosis. In the validation cohorts, WML volumes were analyzed as continuous variables. Within the validation cohorts, we included age, gender, intracranial volume and mean overall protein levels as covariates. Additionally, the time interval between MRI scans and CSF collection was included in the model for the UK Biobank cohort due to the notable time gap.

Linear mixed-effect models including random slope and intercept were used to test the effects of CSF baseline protein levels on WML volume changes and microbleed number variation over time, using protein*time interaction in the model. Age, gender, mean overall protein levels and intracranial volume (for WML only) were used as covariates. To investigate the relationship between protein modules and cSVD, we used linear models to analyze the association between average protein levels of each module and cSVD at baseline. For longitudinal assessments, linear-mixed effect models with random slope and intercept, along with module*time interaction were used to examine the effect of mean protein levels in each module at baseline on the changes of WML volume over time. Longitudinal analyses were not conducted for microbleeds and infarcts due to the limited number of cases showing progression over time, with even fewer cases for specific subtypes of microbleeds and infarcts. To compare the trajectories of key proteins and modules with increasing WML and microbleed pathology, we plotted mean protein levels and mean protein modules against a continuous measure of WML and microbleeds using Generalized Additive Model (GAM). P values for all analyses were adjusted for multiple comparisons using the false discovery rate (FDR) according to the Benjamini-Hochberg procedure, pFDR<0.05.

### Mediation analysis

For the mediation analyses, we investigated the mediation of WML-associated proteins on the association between WML volume (corrected for icv) and executive function or mPACC rate of change. Mediation analyses were conducted using the mediation R package v4.5.0. The significance of the mediating effect was assessed using 1000 bootstrapping iterations and all paths of the mediation model were controlled for age, sex, mean overall protein levels and education. The mPACC composite score was calculated using a mean of z-scores of the Alzheimer’s Disease Assessment Scale (ADAS) delayed recall (weighted double), animal fluency, Mini-Mental State Examination (MMSE), and Symbol Digit Modalities Test (SDMT). The executive function composite score was calculated as the mean of z-scores from the Trail Making Test-A (TMT-A), Trail Making Test-B (TMT-B) and the SDMT, following methodology described previously^112,113^. To calculate the rate of change, linear mixed effect models with random slope and intercept were fitted, with executive function or mPACC as the dependent variable and time in years from the baseline scan as the independent variable. To assess the association between cognitive function (rate of change in executive function or mPACC) and WML-associated proteins at baseline, we used linear models adjusted for age, sex, mean overall protein levels and education.

## Data availability

Pseudonymized data will be shared by request from a qualified academic and as long as data transfer is in agreement with EU legislation on the general data protection regulation and decisions by the Swedish Ethical Review Authority and Region Skåne, which should be regulated in a material transfer agreement.

## Code availability

All code was written using existing packages in R and python and can be accessed by contacting the authors.

## ACKNOWLEDGEMENTS

We would like to acknowledge all the BioFINDER team members as well as participants in the study and their family members for their dedication. The BioFINDER study was supported by the National Institute of Aging (R01AG083740), European Research Council (ADG-101096455), Alzheimer’s Association (ZEN24-1069572, SG-23-1061717), GHR Foundation, Swedish Research Council (2022-00775, 2021-02219, 2018-02052), ERA PerMed (ERAPERMED2021-184), Knut and Alice Wallenberg foundation (2022-0231), Strategic Research Area MultiPark (Multidisciplinary Research in Parkinson’s disease) at Lund University, Swedish Alzheimer Foundation (AF-980907, AF-980832, AF-993465, AF-939981, AF-AF-994229), Swedish Brain Foundation (FO2021-0293, FO2023-0163), Parkinson foundation of Sweden (1412/22), Cure Alzheimer’s fund, WASP and DDLS Joint call for research projects (WASP/DDLS22-066), EU Joint Programme Neurodegenerative Diseases (2019-03401), Rönström Family Foundation (FRS-0003), Konung Gustaf V:s och Drottning Victorias Frimurarestiftelse, Skåne University Hospital Foundation (2020-O000028), Regionalt Forskningsstöd (2022-1259) and Swedish federal government under the ALF agreement (2022-Projekt0080, 2022-Projekt0107). Author JWV is funded through the SciLifeLab & Wallenberg Data Driven Life Science Program (KAW 2020.0239), the Swedish Alzheimer Foundation (AF-994626), and the Crafoord Foundation (20230790). This research has been conducted using the UK Biobank Resource under Application Number 105777. The funding sources had no role in the design and conduct of the study; in the collection, analysis, interpretation of the data; or in the preparation, review, or approval of the manuscript.

## AUTHOR CONTRIBUTIONS

IH, JWV and OH designed the study. IH had full access to the raw data and carried out the final statistical analyses. IH, JWV and OH wrote the manuscript and IH had the final responsibility to submit for publication. SP, ES and OH were responsible for clinical assessment and data collection. CG performed the co-expression module analysis and provided transcriptomics experience. All authors contributed to the interpretation of the results and critically reviewed the manuscript.

