## Supplementary Figures for "Identification of distinct and shared biomarker panels in different manifestations of cerebral small vessel disease through proteomic profiling"

### Supplementary Fig. 1. Distribution of WML values and threshold for WML positivity

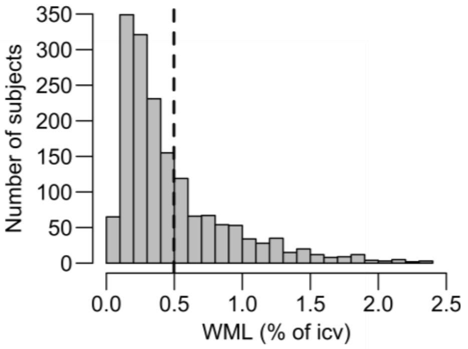


Legend: (a) Histogram depicting the distribution of WML values expressed as a percentage of intracranial volume (icv). A dashed line at 0.497 indicates the threshold for WML positivity. Participants in the lower two tertiles do not exhibit significant WML pathology, whereas those in the upper tertile are classified as exhibiting WML pathology.

**Supplementary Fig. 2. Pathway analysis of common upregulated CVD-associated proteins**

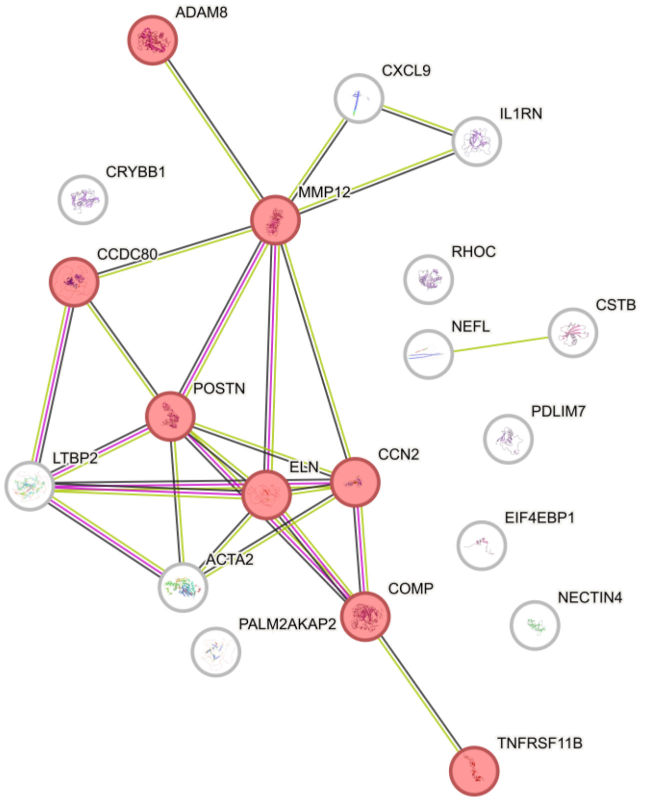


Legend: (a) Protein-protein interactions were identified using the STRING software with a minimum interaction score threshold of >0.4. Proteins highlighted in red are involved in extracellular matrix organization, a biological process that is significantly enriched (pFDR < 0.05) according to Gene Ontology (GO) analysis.

**Supplementary Fig. 3. Progression of WML pathology**

A

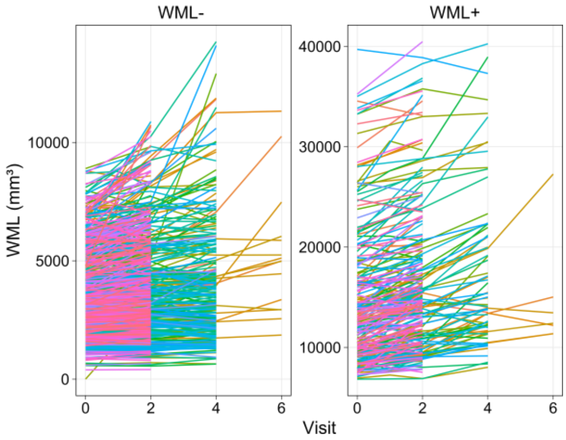


Legend: (a) Spaghetti plot illustrating the progression of WML volumes across successive visits. Individual trajectories represent WML volume change for each participant. For clarity, the plots are divided: the left panel (WML-) depicts subjects without WML pathology, while the right panel (WML+) shows subjects with WML pathology.
