## Extended data for "Identification of distinct and shared biomarker panels in different manifestations of cerebral small vessel disease through proteomic profiling"

### Extended Data Figures and Tables

#### Extended Data Fig. 1. Overview of the study design

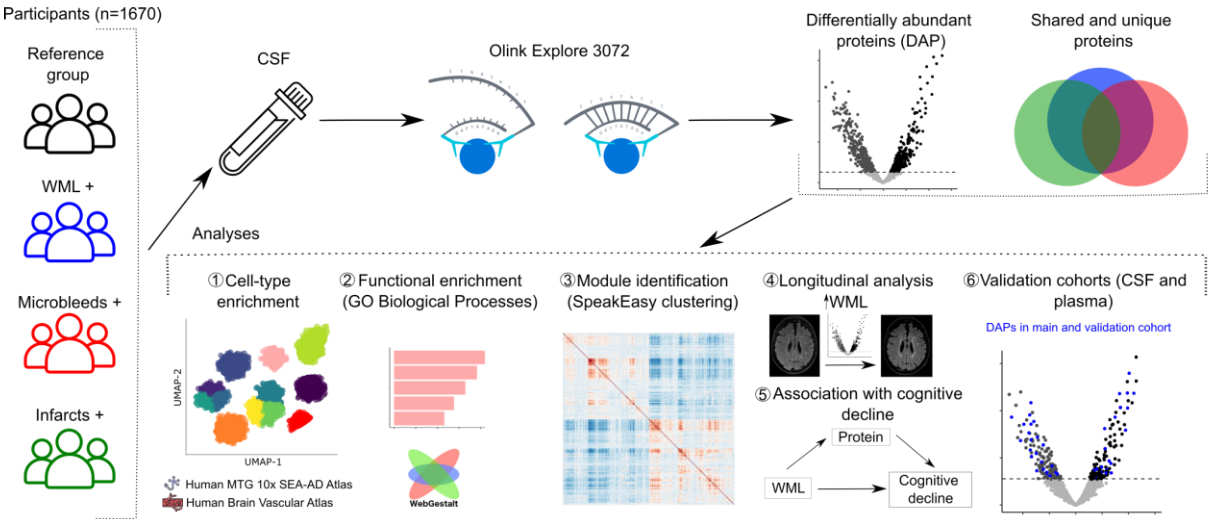


Legend: Cerebrospinal fluid (CSF) protein levels were quantified using Olink proteomics antibody-based Proximity Extension Assay (PEA) technology in 1670 participants from the BioFINDER-2 study. We analyzed differential protein abundance in CSF across various cSVD manifestations, namely white matter lesions (WML), microbleeds, and infarcts. Subsequently, for proteins identified as differentially abundant, cell-type specific enrichment was assessed using the Expression Weighted Cell Type Enrichment (EWCE) approach (1). Additionally, functional enrichment was performed utilizing Gene Ontology (GO) databases (2). Protein co-abundance modules were identified using a clustering algorithm, to explore the association between these biological modules and cSVD manifestations (3). Longitudinal analyses were performed to evaluate the association of Olink proteins with WML or microbleed progression (4). Mediation analysis was conducted to identify whether WML-associated proteins mediate the relationship between WML and cognitive decline (5). Validation was performed in the same cohort using plasma, and in three distinct cohorts using CSF and plasma samples, and their utility in predicting future cerebrovascular events was assessed (6).
Abbreviations: WML=White Matter Lesions, CSF=Cerebrospinal Fluid, GO=Gene Ontology.

### Extended Data Table 1. BioFINDER-2 cohort demographics

| | White matter lesions | | Microbleeds | | Infarcts | |
|---|---|---|---|---|---|---|
| | **WML- (N=1113)** | **WML+ (N=557)** | **MB- (N=1350)** | **MB+ (N=269)** | **Infarcts- (N=1445)** | **Infarcts+ (N=225)** |
| **Age (y)** | | | | | | |
| Mean (SD) | 65.5 (±12.7) | 75.7 (±6.03)[c] | 67.7 (±12.5) | 74.3 (±6.99)[c] | 67.9 (±12.4) | 75.2 (±6.41)[c] |
| **Sex** | | | | | | |
| Male | 505 (45 %) | 302 (54 %)[c] | 629 (47 %) | 157 (58 %)[c] | 673 (47 %) | 134 (60 %)[c] |
| Female | 608 (55 %) | 255 (46 %)[c] | 721 (53 %) | 112 (42 %)[c] | 772 (53 %) | 91 (40 %)[c] |
| ***APOE* ε4 positivity[1] (%)** | 551 (50 %) | 251 (45 %) | 643 (48 %) | 136 (51 %) | 711 (49 %) | 91 (40 %)[a] |
| **Education[2] (y)** | | | | | | |
| Mean (SD) | 13.1 (±3.60) | 12.0 (±3.75)[c] | 12.8 (±3.66) | 12.6 (±3.85) | 12.8 (±3.64) | 12.1 (±3.92)[b] |
| **MMSE, baseline score** | | | | | | |
| Mean (SD) | 27.5 (±3.32) | 26.0 (±4.03)[c] | 27.3 (±3.45) | 25.8 (±4.12)[c] | 27.1 (±3.62) | 26.3 (±3.76)[c] |
| **Cognitive status** | | | | | | |
| Dementia | 147 (13 %) | 165 (30 %)[c] | 210 (16 %) | 88 (33 %)[c] | 248 (17 %) | 64 (28 %)[c] |
| MCI | 262 (24 %) | 194 (35 %)[c] | 344 (25 %) | 95 (35 %)[b] | 376 (26 %) | 80 (36 %)[b] |
| CU | 704 (63 %) | 198 (36 %)[c] | 796 (59 %) | 86 (32 %)[c] | 821 (57 %) | 81 (36 %)[c] |
| **AD diagnosis** | 138 (12 %) | 134 (24 %)[c] | 189 (14 %) | 72 (27 %)[c] | 227 (16 %) | 45 (20 %) |
| **PD diagnosis** | 62 (6 %) | 14 (3 %)[b] | 62 (5 %) | 9 (3 %) | 69 (5 %) | 7 (3 %) |
| **WML (% of ICV)** | | | | | | |
| Mean (SD) | 0.26 (±0.11) | 0.94 (±0.40)[c] | 0.42 (±0.36) | 0.79 (±0.51)[c] | 0.44 (±0.38) | 0.78 (±0.45)[c] |
| **≥ Microbleeds, n present (%)** | 97 (9 %) | 172 (31 %)[c] | 0 (0 %) | 269 (100%) | 193 (13 %) | 76 (34 %)[c] |
| **≥ Microbleed location, n present lobar (%)** | 72 (6 %) | 137 (25 %)[c] | 0 (0 %) | 209 (78 %) | 151 (10 %) | 58 (26 %)[c] |
| **≥ Microbleed location, n present deep (%)** | 22 (2 %) | 71 (13 %)[c] | 0 (0 %) | 93 (35 %) | 52 (4 %) | 41 (18 %)[c] |
| **≥ Infarcts, n present (%)** | 75 (7 %) | 150 (27 %)[c] | 139 (10 %) | 76 (28 %)[c] | 0 (0 %) | 225 (100 %) |
| **≥ Subcortical infarcts/lacunes, n present (%)** | 46 (4 %) | 92 (17 %)[c] | 88 (7 %) | 46 (17 %)[c] | 0 (0 %) | 138 (61 %) |
| **≥ Cortical and cerebellar infarcts, n present (%)** | 36 (3 %) | 76 (14 %)[c] | 64 (5 %) | 41 (15 %)[c] | 0 (0 %) | 112 (50 %) |
| **Cardiovascular disease, n yes (%)** | 452 (41 %) | 342 (61 %)[c] | 601 (45 %) | 161 (60 %)[c] | 639 (44 %) | 155 (69 %)[c] |
| **Hypertension, n yes (%)** | 345 (31 %) | 266 (48 %)[c] | 466 (35 %) | 122 (45 %)[b] | 485 (34 %) | 126 (56 %)[c] |
| **Ischemic heart disease, n yes (%)** | 71 (6 %) | 70 (13 %)[c] | 100 (7 %) | 32 (12 %)[a] | 107 (7 %) | 34 (15 %)[c] |
| **Stroke/TIA, n yes (%)** | 55 (5 %) | 77 (14 %)[c] | 87 (6 %) | 42 (16 %)[c] | 68 (5 %) | 64 (28 %)[c] |
| **Atrial fibrillation, n yes (%)** | 21 (2 %) | 18 (3 %) | 22 (2 %) | 14 (5 %)[c] | 31 (2 %) | 8 (4 %) |
| **Diabetes, n yes (%)** | 102 (9 %) | 88 (16 %)[c] | 146 (11 %) | 38 (14 %) | 149 (10 %) | 41 (18 %)[c] |
| **Dyslipidemia, n yes (%)** | 297 (27 %) | 257 (46 %)[c] | 404 (30 %) | 125 (46 %)[c] | 427 (30 %) | 127 (56 %)[c] |

Data are shown as mean ± standard deviation, unless specified otherwise. Information for microbleeds was missing for n=51 participants.

Abbreviations: y=years, *APOE*ε4=apolipoprotein E genotype (carrying at least one ε4 allele); MMSE=Mini-Mental State Examination; MCI=Mild Cognitive Impairment; CU=Cognitively Unimpaired; AD=Alzheimer's disease; PD=Parkinson's disease; WML=White Matter Lesions, ICV=Intra Cranial Volume; TIA=Transient ischemic attack

[1]: APOE status was missing for 147 individuals.

[2]: Education information was missing for 31 individuals.

Statistical significance: [a] $p<0.05$, [b] $p<0.01$, [c] $p<0.001$

**Extended Data Fig. 2. Differential protein expression when models corrected for AD and PD diagnosis**

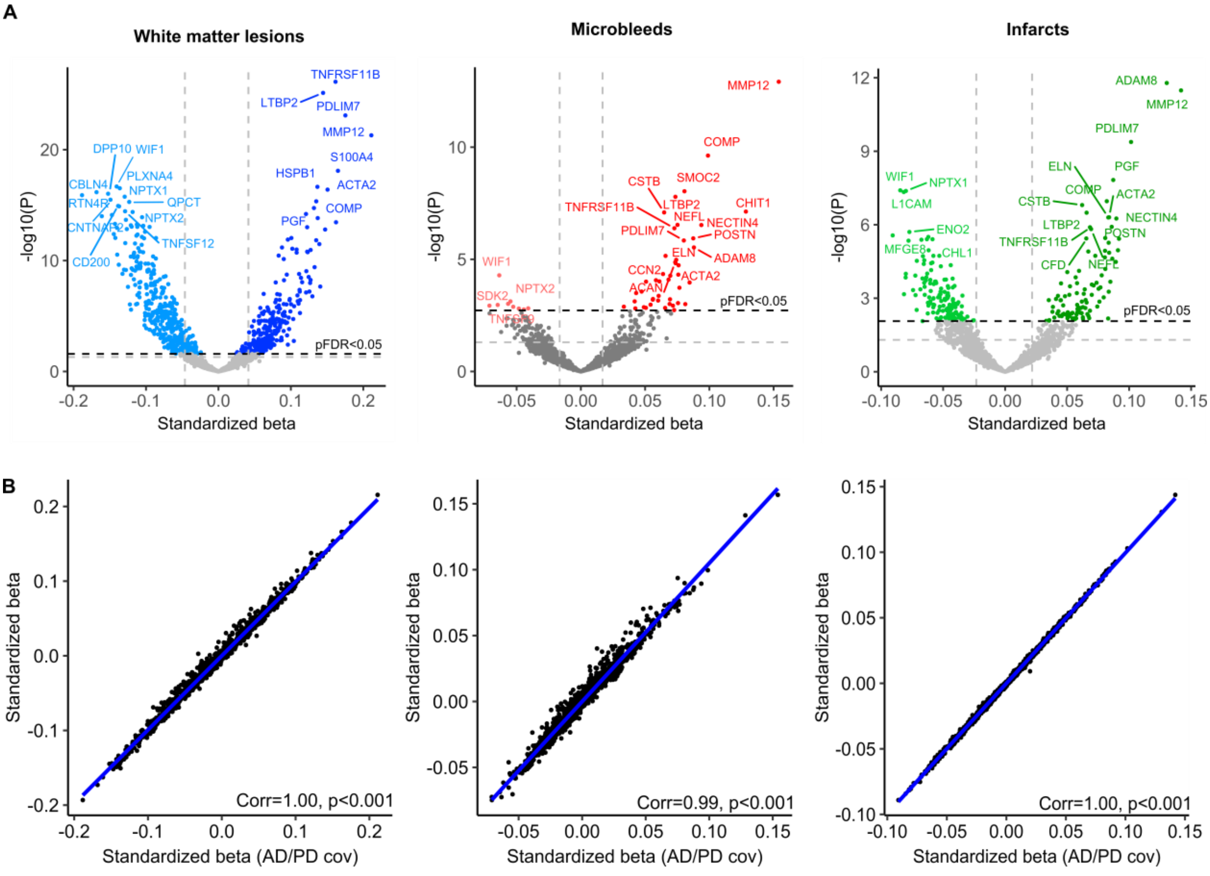


Legend: (a) Volcano plot showing differentially abundant proteins (DAPs) when comparing individuals with and without a specific cSVD pathology while correcting for AD and PD diagnosis. The models were adjusted for age, sex, and average protein level. The dashed lines represent significance threshold at α=0.05 before (gray) and after (black) FDR correction. Proteins below the p[FDR]<0.05 threshold were considered significant. For clarity, only the top 20 proteins are labeled. (b) Comparison of the standardized betas from the main analysis assessing differential protein abundance vs. the standardized betas when further adding AD and PD diagnosis as covariates.

**Extended Data Fig. 3. Characterization of proteomic profiles across different microbleed and infarct types**

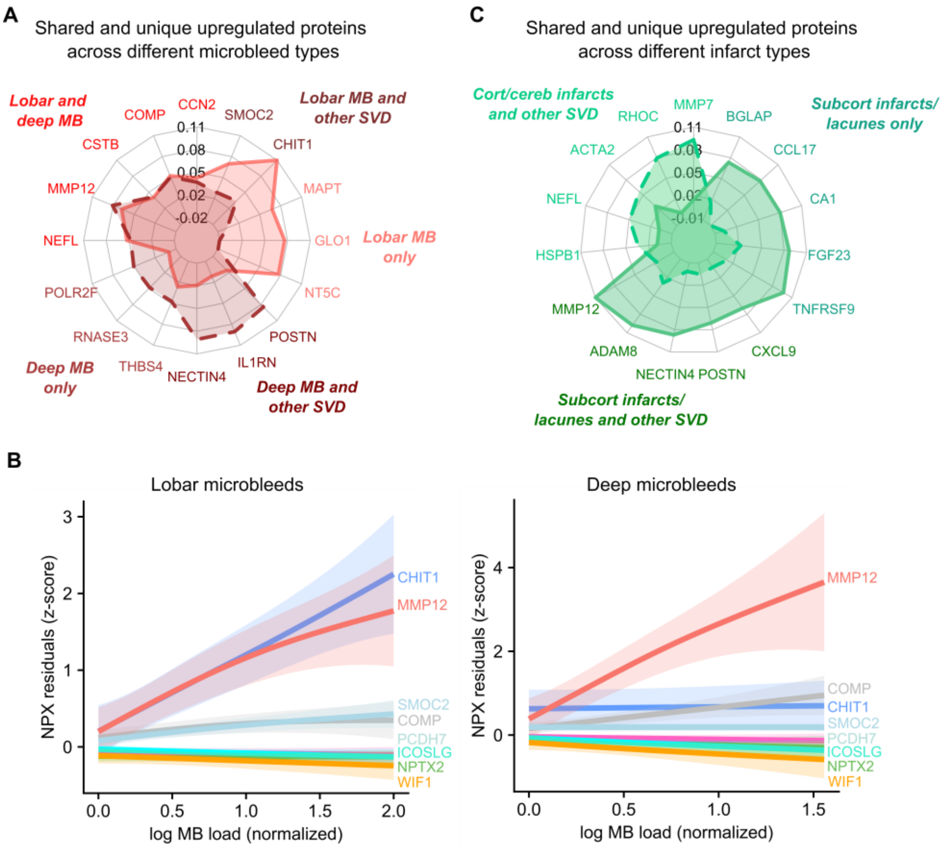


Legend: (a) Radar chart displaying standardized beta coefficients for top proteins across various microbleed types. Proteins shown may be either shared across different SVD subtypes, or uniquely upregulated within microbleed subtypes. Each axis represents a different protein, with axis length indicating the magnitude of its standardized beta coefficient. (b) Top proteins associated with microbleeds (Figure 1g), plotted against normalized lobar microbleed (a) and deep microbleed load (b). Shaded areas correspond to the 95% confidence interval. (c) Radar chart displaying standardized beta coefficients for top proteins across various infarct types. Proteins shown may be either shared across different SVD subtypes, or uniquely upregulated within infarct subtypes. Each axis represents a different protein, with axis length indicating the magnitude of its standardized beta coefficient.

**Extended Data Fig. 4. Clustering algorithm derived modules of protein co-abundance and their association with cSVD**

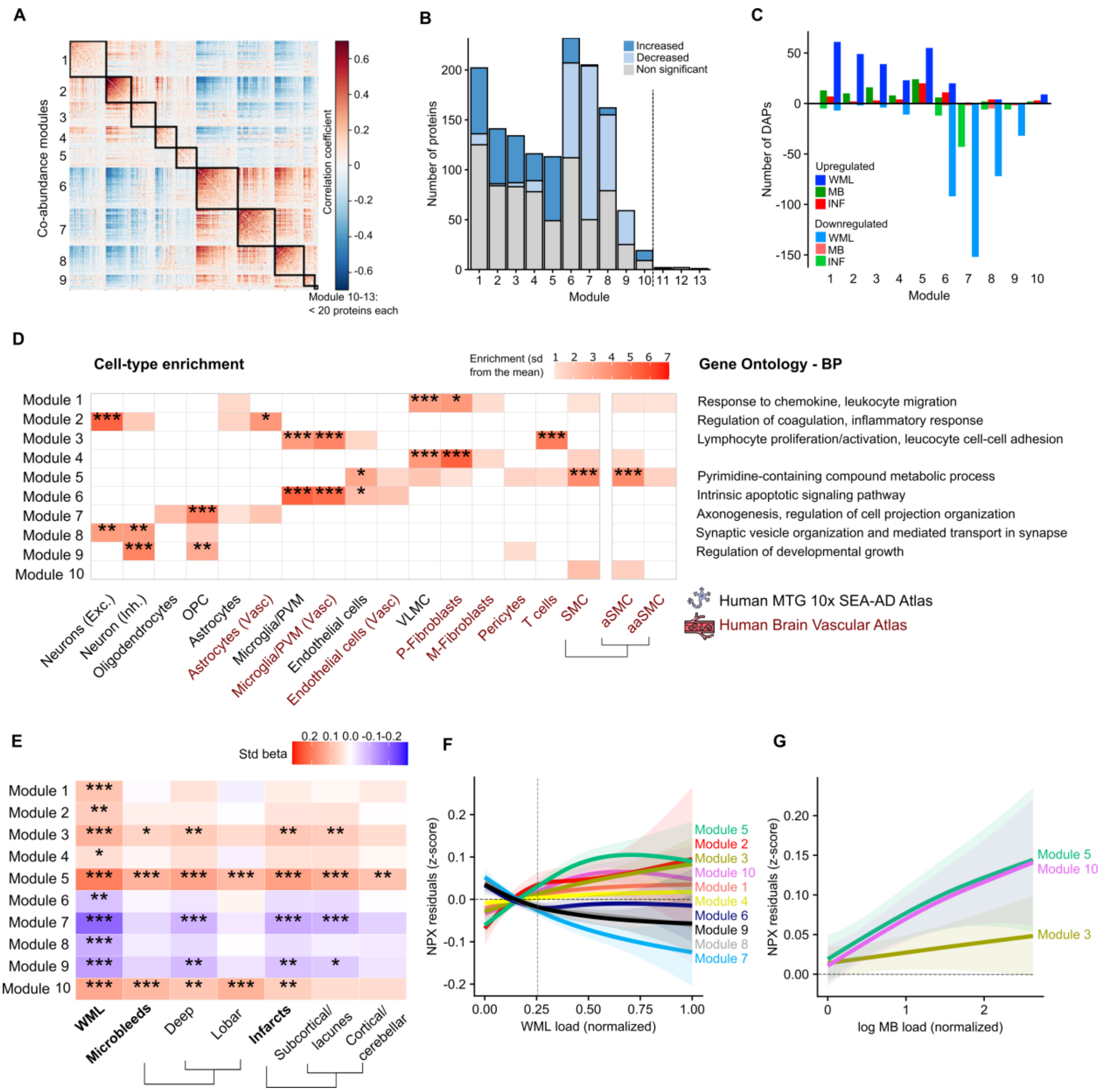


Legend: (a) Heatmap depicting protein-protein correlation matrix within the Olink data. Black outlined boxes indicate co-abundance modules, with modules 10-13 comprising fewer than 20 proteins each. (b) Distribution of proteins across co-abundance modules. Bar heights represent the number of proteins in each module, categorized by changes in abundance in all three cSVD manifestations combined: increased (dark blue), decreased (pale blue), and non-significant (gray). Modules 11 to 13, demarcated by a dashed line, contain few proteins, and are not used in subsequent analysis. (c) Bar chart illustrating the module-wise distribution of DAP across various cSVD conditions. Upregulated proteins are represented above the horizontal axis, and downregulated proteins are shown below the axis in corresponding colors for each condition. (d) Heatmap displaying cell-type enrichment across protein co-abundance modules, using single-cell transcriptomics data from the Human MTG 10x SEA-AD and the Human Brain Vascular Atlas. Adjacent to the heatmap, key biological processes identified through Gene Ontology (GO) – Biological Processes (BP) enrichment analysis are listed for each module. (e) Heatmap of standardized beta coefficients illustrating associations between biological modules and WML, microbleeds and infarcts. (f, g), Trajectories of average protein expression within significant modules as a function of normalized (f) WML load and (g) microbleed load, showcasing the relationship between protein abundance and disease burden. Shaded areas correspond to the 95% confidence interval. The vertical line indicates the cutoff for WML positivity.

*corresponds to $p_{FDR}<0.05$, **corresponds to $p_{FDR}<0.01$, ***corresponds to $p_{FDR}<0.001$

**Extended Data Fig. 5. Characterization of WML-associated proteins mediating the relationship between WML and cognitive decline as measured by mPACC**

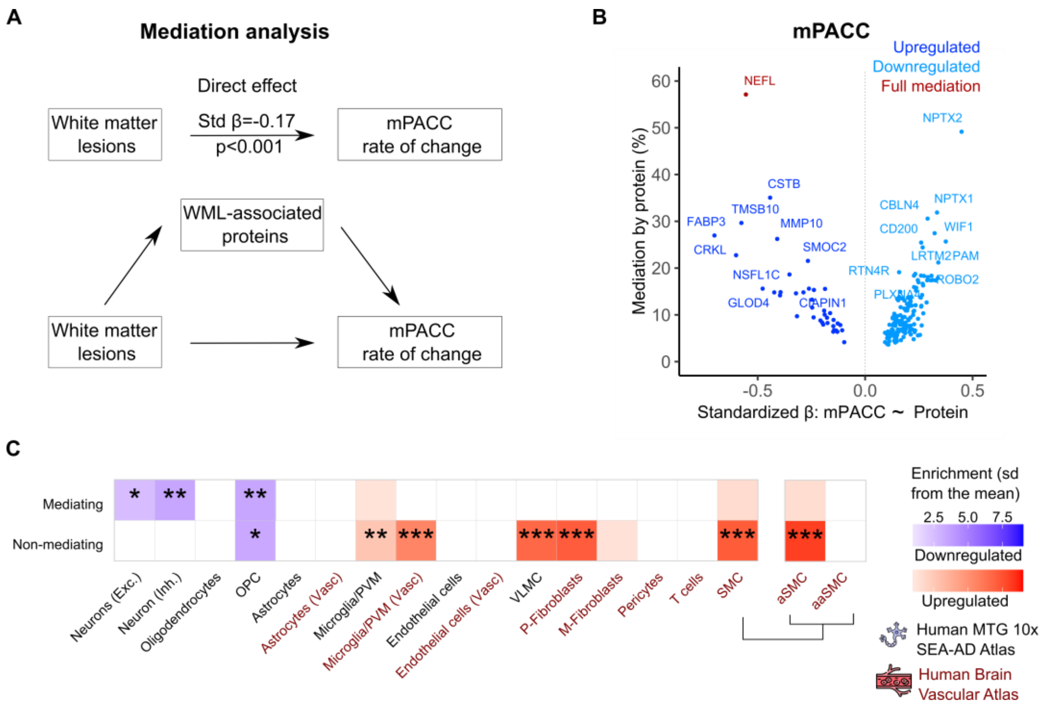


Legend: (a) Path diagram of the mediation analysis models. (b) Volcano plots showing the extent to which WML-associated proteins mediate the association between WML and mPACC rate of change by the standardized beta values derived from the association between proteins and mPACC rate of change. The top 10 upregulated and top 10 downregulated proteins with highest mediation effect are labeled. (c) Comparison for cell-type enrichment between WML-associated proteins that mediate and those that do not mediate the interaction between WML and mPACC rate of change based on single cell transcriptomics data from the MTG 10x SEA-AD Atlas and the Human Brain Vascular Atlas using the EWCE package. For this analysis, the 1331 Olink proteins were used as background.

*corresponds to $p_{FDR}<0.05$, **corresponds to $p_{FDR}<0.01$, ***corresponds to $p_{FDR}<0.001$.

Abbreviations: OPC = oligodendrocyte precursor cells, PVM = perivascular macrophages, VLMC = vascular leptomeningeal cell, P-Fibroblasts=perivascular fibroblasts, M-Fibroblasts = meningeal fibroblasts, SMC = smooth muscle cell, aSMC = arterial smooth muscle cell, aaSMC=arteriolar smooth muscle cell

**Extended Data Table 2. Demographics of validation cohorts: BioFINDER-1, ADNI and UK Biobank**

| | BioFINDER-1 (n=383) | ADNI (n=729) | UK Biobank (N=5712) |
|---|---|---|---|
| **Age (y)** | 73.3 (± 5.02) | 73.4 (± 7.45) | 54.2 (± 7.75) |
| **Sex** | | | |
| Male | 158 (41%) | 414 (57%) | 3067 (54%) |
| Female | 225 (59%) | 315 (43%) | 2645 (46%) |
| **Education (y)**[1] | 12.1 (± 3.64) | 15.9 (± 2.85) | 12.2 (± 2.41) |
| ***APOE* ε4 positivity (%)**[2] | | | |
| No | 219 (57%) | 368 (50%) | 1318 (23%) |
| Yes | 130 (34%) | 361 (50%) | 3533 (62%) |
| **MMSE, baseline score** | 28.6 (± 1.42) | 27.1 (± 2.64) | |
| **Cognitive status** | | | |
| CU | 319 (83%) | 171 (23%) | |
| MCI | 64 (17%) | 415 (57%) | |
| Dementia | 0 (0%) | 143 (20%) | |
| **WML (% of ICV)** | 0.317 (± 0.269) | 0.385 (± 0.393) | 0.33 (± 0.45) |

Data are shown as mean ± standard deviation, unless specified otherwise.
Abbreviations: y=years, SD=standard deviation; *APOE*ε4=apolipoprotein E genotype (carrying at least one ε4 allele), MMSE=Mini-Mental State Examination; WML=White Matter Lesions, ICV=Intra Cranial Volume.

[1]: Education information was missing for 2 individuals in the BioFINDER-1 cohort and for 2739 individuals in the UK Biobank cohort.

[2]: APOE status was missing for 34 individuals in the BioFINDER-1 cohort and for 861 individuals in the UK Biobank cohort.

**Extended Data Fig. 6. Differential protein expression related to continuous measures of WML volume**

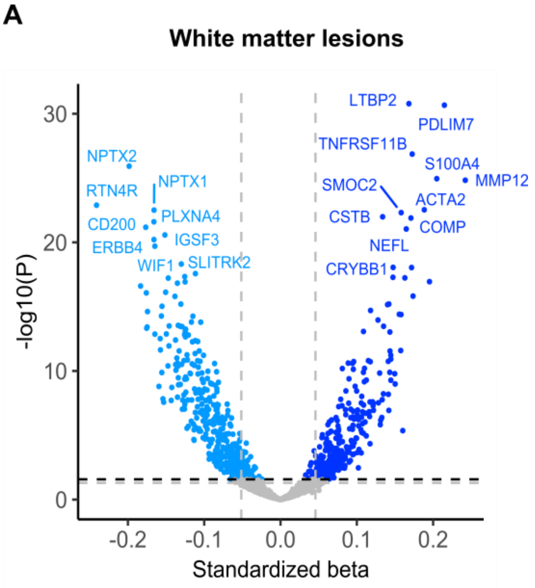


Legend: (a) Volcano plot showing differentially abundant proteins (DAPs), analyzed with WML as a continuous variable, and adjusting for age, sex, intracranial volume, and average protein level. The dashed lines represent significance threshold at α=0.05 before (gray) and after (black) FDR correction. Proteins below the p[FDR]<0.05 threshold were considered significant. For clarity, only the top 20 proteins are labeled.

**Extended Data Figure 7. Validation of DAP in subjects with WML in the UK Biobank plasma dataset**

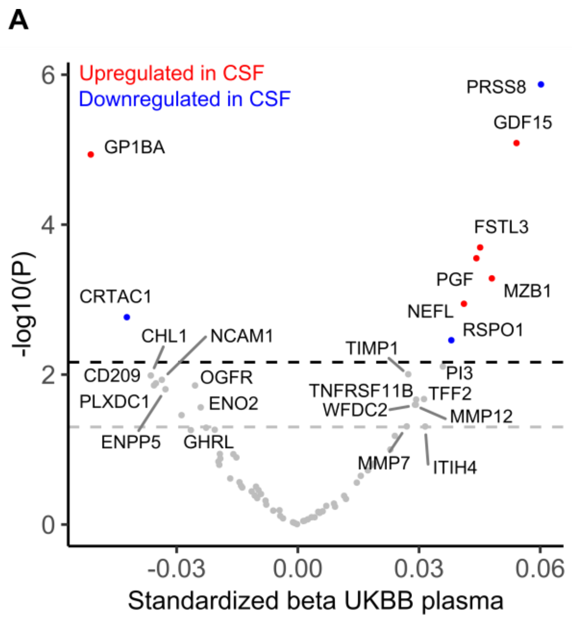


Legend: **(a)** Volcano plots showing differentially abundant proteins (DAP) when comparing individuals with and without WML in the UK Biobank. The models are adjusted for age, sex, average protein level and time interval between MRI scans and CSF collection. The dashed lines represent significance threshold at α=0.05 before (gray) and after (black) FDR correction. Proteins below the p[FDR]<0.05 threshold were considered significant. For clarity, only the top 20 proteins are labeled.

**Extended Data Table 3. Demographics of the train and test set in UK Biobank validation cohort**

| | Train set (any cerebrovascular disease, n=40205) | Train set (infarcts, n=41159) | Test set (n=10447) |
|---|---|---|---|
| **Age (y)** | 56.6 (± 8.19) | 56.7 (± 8.20) | 56.6 (± 8.27) |
| **Sex** | | | |
| Male | 21884 (54 %) | 22339 (54 %) | 5624 (54 %) |
| Female | 18321 (46 %) | 18820 (46 %) | 4823 (46 %) |
| **Systolic blood pressure**[1] | 137 (± 18.5) | 138 (± 18.6) | 138 (± 18.7) |
| **Diastolic blood pressure**[1] | 82.1 (± 10.1) | 82.1 (± 10.1) | 82.2 (± 10.2) |
| **Glucose (mg/ml)**[2] | 92.5 (± 22.6) | 92.6 (± 22.7) | 92.7 (± 22.8) |
| **Total cholesterol (mg/ml)**[3] | 219 (± 44.5) | 219 (± 44.6) | 218 (± 45.0) |
| **Body mass index (BMI)**[4] | 27.4 (± 4.79) | 27.4 (± 4.79) | 27.5 (± 4.81) |
| **Atrial fibrillation, n yes (%)** | 607 (2 %) | 639 (2 %) | 168 (2 %) |
| **Diabetes, n yes (%)** | 934 (2 %) | 999 (2 %) | 238 (2 %) |
| **Personal or family history of disease, n yes (%)** | 1040 (3 %) | 1110 (3 %) | 289 (3 %) |
| **Current tobacco smoking, n yes (%)**[5] | 3048 (8 %) | 3143 (8 %) | 814 (8 %) |
| **With cerebrovascular event** | 441 | 441 | 111 |
| **With infarct** | 183 | 183 | 46 |

Data are shown as mean ± standard deviation, unless specified otherwise.

[1]: Systolic and diastolic blood pressure information was missing for 4309 individuals in the UK Biobank cohort.
[2]: Glucose levels information was missing for 6703 individuals.
[3]: Total cholesterol information was missing for 2520 individuals.
[4]: BMI information was missing for 238 individuals.
[5]: Current tobacco smoking was missing for 59 individuals.

**Extended Data Figure 8. Shapley Additive exPlanations (SHAP) feature importance for cerebrovascular events and infarct prediction using Random forest classifier model.**

A

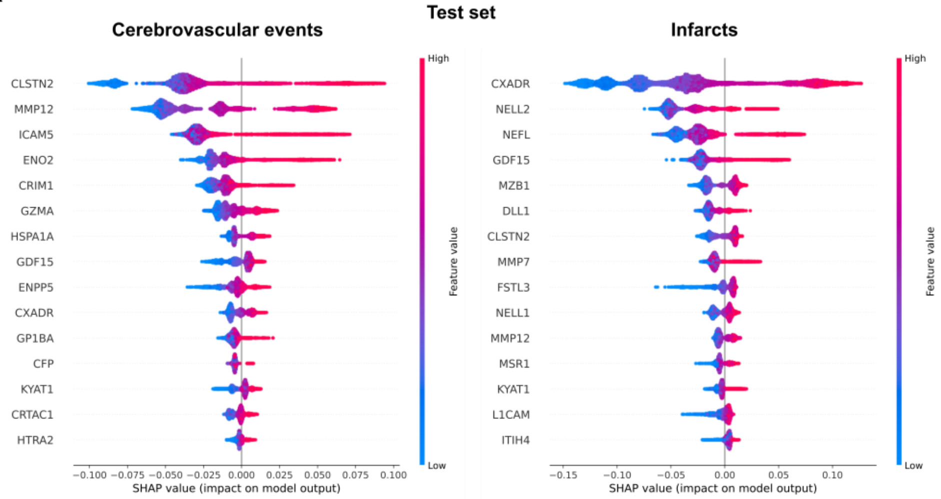


Legend: (a) SHapley Additive exPlanations (SHAP) feature importance for the model incorporating age, sex, clinical variables, proteins and average protein level for (a) cerebrovascular events and (b) infarcts. For visualization purposes, we included only the top 15 features, shown in order of importance, from most important (top) to less important (bottom).
